# How many large language models does title–abstract screening need? Theory, two replications and an evaluation at real prevalence

**DOI:** 10.64898/2026.11.25.26369001

**Authors:** Eduardo Quiñelen, Ignacio Neumann, Miguel Contreras

**Affiliations:** Department of Health Technology Assessment and Evidence-Based Health, Ministry of Health of Chile, Santiago, Chile; Department of Digital Transformation, Ministry of Health of Chile, Santiago, Chile

**Keywords:** Systematic reviews, Study selection, Large language models, Artificial intelligence, Ensemble methods, Automation, Evidence synthesis

## Abstract

**Background and Objectives:** Researchers increasingly combine large language models (LLMs) to screen titles and abstracts, assuming that more models make screening safer. We estimated how many models a screening ensemble needs, given correlated errors, and tested the answer on independent reviews and at real prevalence.

**Methods:** We modelled correlated votes with a beta-binomial model and estimated, by bootstrap, the smallest ensemble capturing 95% of the net-benefit gain attainable over the best single model, accounting for human reading time and machine cost. Eleven LLMs from four developers voted blind on records from 13 SYNERGY reviews (derivation). We tested predictions fixed in advance on 10 and 16 Cochrane reviews from a public screening benchmark (replications) and evaluated a two-voter pipeline with an adjudicator on 11 complete benchmark collections and six reviews published in 2026.

**Results:** Errors on relevant records were correlated (intraclass correlation 0.56 among eight different models; 0.75–0.78 among roles or copies of one model): eight models carried the information of about 1.6 independent ones. In derivation, the best single model, pair and trio reached sensitivities of 0.920, 0.975 and 0.981. Three or fewer models captured 95% of the attainable gain in at least 86%, 88% and 80.8% of bootstrap replicates of every scenario in the derivation set and the two replications, respectively. Six of seven registered predictions held in replication 1 and three of five sealed predictions in replication 2, where a fourth model added 2.1 percentage points to trios on average but 0.3 to the best trio. At real prevalence, the pipeline sent 94 of 96 included studies to human reading in the benchmark collections and 163 of 178 in the 2026 reviews (12.1% of 12,196 records sent to reading); its more sensitive voter alone sent 259 of 274 with 12.0%, at under a quarter of the machine cost.

**Conclusion:** Because LLM errors are correlated, our results suggest two or three well-chosen models may suffice, but this needs deeper exploration in future studies. Which models are combined and how matter as much as how many, and any ensemble should be compared with its best single member.

**Plain Language Summary:** People who summarise medical research must read thousands of short article summaries to find the few studies that matter. Artificial intelligence models can help, and researchers increasingly combine several models in the hope of missing fewer studies. We found that these models tend to make the same mistakes, so adding more of them helps little: two or three well-chosen models were nearly as good as any larger group, though this needs more study. We wrote our predictions down before testing them on new sets of reviews, including reviews published after the main model’s training data were collected. In 17 real reviews, a group of three models passed 94% of the relevant studies to a human reader, who would have had to read about one in eight summaries. One of its two voting models alone did about as well for under a quarter of the price. Teams should check whether a group of models beats its best member before paying for more.

**What is new?:** *Key findings:* Errors of large language models in title–abstract screening were highly correlated: eight different models carried the information of about 1.6 independent ones. - The best pair or trio captured 95% of the attainable gain in most bootstrap replicates of a derivation set and two replications with predictions fixed in advance. - At real prevalence, a three-model pipeline and its more sensitive voter alone sent almost the same number of included studies to human reading (257 and 259 of 274).

*What this adds to what is known:* Earlier studies compared ensemble sizes or aggregation rules; this study derives the size from error correlation, reading time and machine cost, and tests it on independent reviews.

*What is the implication and what should change now:* Our results suggest that review teams may not need more than three models, although this needs confirmation; they should measure candidate models locally through the route they will use, and keep an ensemble only if it beats its best single member.

## 1. Introduction

Title and abstract screening is the most labour-intensive step of a systematic review. Standards prefer two independent reviewers [1] because a single reviewer misses a non-negligible fraction of eligible studies: 13% in a randomised trial [2], and 5%–9% in other estimates [3,4]. Large language models (LLMs) are now evaluated as a second reviewer, as a filter or as autonomous screeners [5–8], and a growing number of workflows combine several models and aggregate their votes [9–14].

Ensembles implicitly rely on Condorcet’s jury theorem [15]: if voters are independent and each is right more often than not, a larger jury is more often right. LLMs are not independent. Their errors coincide, more so within a developer or architecture, but also across developers [16,17]. Dependence reduces, and can cancel, the benefit of a larger jury [18–20]; dependent diagnostic tests likewise gain less when combined [21]. Outside health, nine LLM judges were worth about two independent votes [22], and error correlation can change the optimal voting threshold of LLM committees [23].

Previous studies compared ensemble sizes, aggregation rules or model pairs [9–14]. To our knowledge, none derived the number of models a screening ensemble needs from error correlation, the reading time created by false positives and machine cost, and then tested it on independent reviews with predictions fixed in advance. We asked how many models, and which, a screening ensemble needs, and tested the answer in two replications and at real prevalence.

## 2. Methods

### 2.1 Design and reporting

This methodological study had six stages (Fig. 1; Table 1): a theoretical model fitted to a calibration corpus; an exploratory derivation study; two replications on independent reviews; and two evaluations of a screening pipeline at real prevalence. In the derivation and replications, every model voted on every record blind to the reference standard and to other votes, so all subsets and aggregation rules came from the same votes and comparisons are paired. In the derivation, the corpus and eligibility protocols were frozen before screening, but the analysis plan was not registered. From replication 1 onwards, the corpus, protocols and predictions with their decision rules (from replication 2, also the analysis scripts) were frozen and hashed (SHA-256) before any vote, with local but no external time-stamps; replication 1 was also registered on the Open Science Framework (OSF) before its data were downloaded [24]. Deviations are listed in Appendix C. Reporting follows the applicable items of STARD 2015 [25] and TRIPOD-LLM [26] (Appendices F and G).

**Fig. 1.**
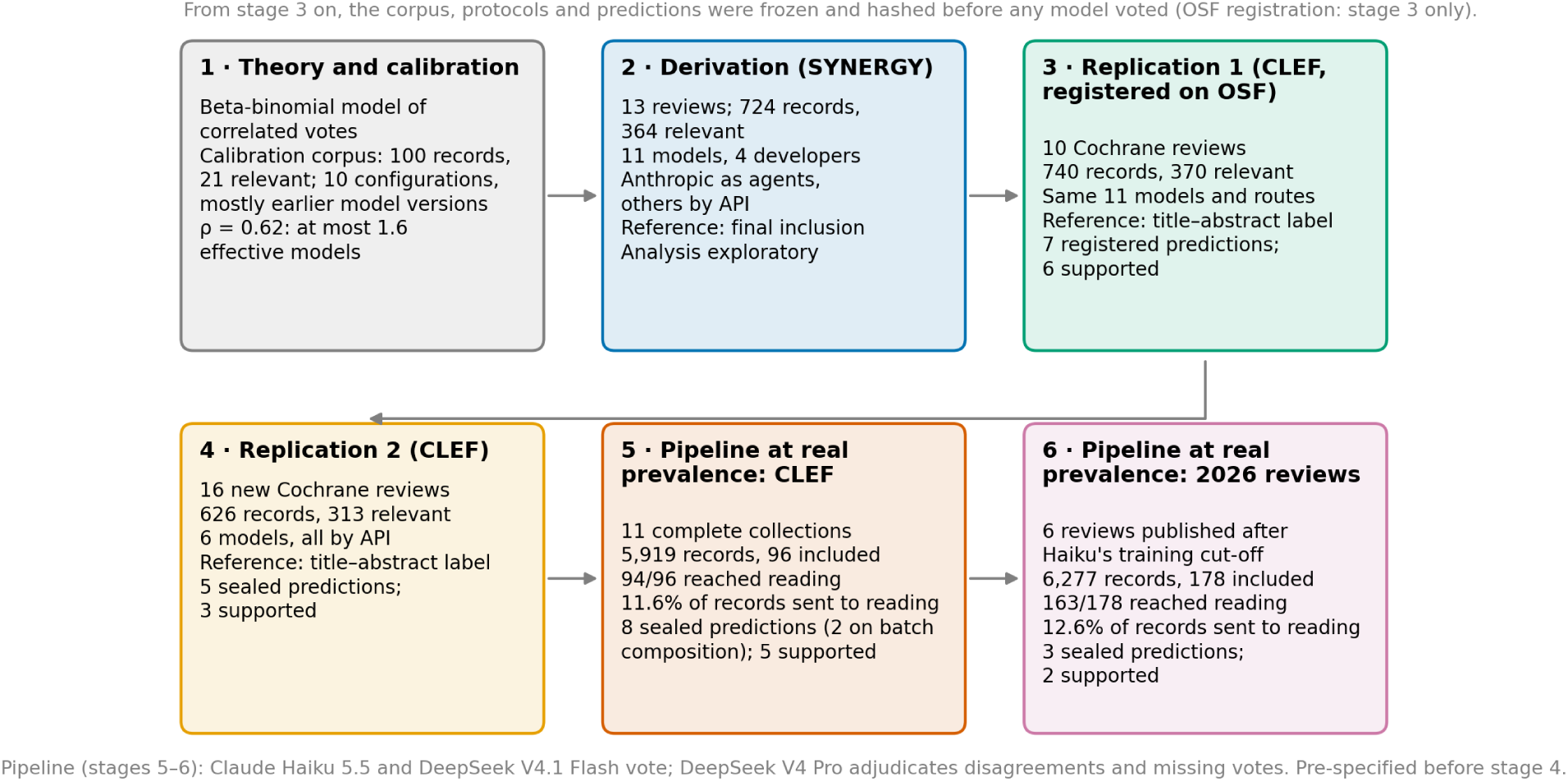
Stages of the study. Each stage answered a question left open by the previous one. From replication 1 onwards, the corpus, protocols and predictions of each new data set were frozen and hashed before any model voted; in the derivation only the corpus and protocols were frozen, and its analysis was exploratory. The pipeline evaluated at real prevalence was pre-specified before replication 2.

**Table 1.** Data sets, models and reference standards of each stage.

| Stage | Source and selection | Reviews (DTA/INT) | Records | Relevant | Reference standard | Models (route) | Pre-specification |
|---|---|---|---|---|---|---|---|
| Calibration | One review of biologics for severe asthma | 1 | 100 | 21 | Human screening decision | 10 configurations, mostly earlier versions (Anthropic, Google) | Protocol frozen; analysis post hoc |
| Derivation | SYNERGY v1.0; health reviews with recoverable criteria; relevant records up to 45 plus an equal random number of irrelevant ones | 13 | 724 | 364 | Final inclusion | 11 (Anthropic as agents; others by API) | Corpus and protocols frozen; analysis exploratory |
| Replication 1 | CLEF TAR 2019; reviews with 10–150 relevant records in registered pseudo-random order; relevant records up to 45 plus an equal random number | 10 (5/5) | 740 | 370 | Title–abstract label | 11 (as above) | OSF registration (7 predictions) |
| Replication 2 | CLEF TAR 2019; new reviews; relevant records up to 20 plus an equal random number | 16 (8/8) | 626 | 313 | Title–abstract label | 6 (all by API) | Local seal, 28 Oct 2026 (5 predictions) |
| Real prevalence, CLEF | CLEF TAR 2019; complete collections, no sampling | 11 (6/5) | 5,919 | 217 (96 included) | Final inclusion; title–abstract label | Pipeline (by API) | Local seals, 28–29 Oct 2026 (8 predictions, 2 on batch composition) |
| Real prevalence, 2026 | Reviews published 1 Jul–30 Oct 2026 with open licences; searches rebuilt | 6 (2/4) | 6,277 | 178 included | Final inclusion | Pipeline (by API) | Local seal, 1 Nov 2026 (3 predictions) |

DTA, diagnostic test accuracy; INT, intervention; API, application programming interface; OSF, Open Science Framework; CLEF TAR, Conference and Labs of the Evaluation Forum Technology Assisted Reviews. Relevant: relevant at title–abstract level in the CLEF samples; finally included in SYNERGY and in the 2026 reviews (in the 2026 reviews, PubMed records of included studies; a study may have several records). Pipeline: Claude Haiku 5.5 and DeepSeek V4.1 Flash vote on every record; DeepSeek V4 Pro adjudicates disagreements and records with one missing vote.

### 2.2 Theoretical model

We treat the votes of K models on a record as exchangeable within its true class, with a record-level inclusion probability that follows a Beta distribution; the intraclass correlation ρ then measures how much model errors coincide. Three results follow (proofs in Appendix A). (i) The effective number of independent models is K/[1 + (K − 1)ρ] and never exceeds 1/ρ. (ii) Under the rule “send the record to a human if any model includes it”, missed relevant records fall as a power of K, not geometrically. (iii) Calling a third model only when two disagree gives, record by record, the decisions of a majority of three. We fitted the model by maximum likelihood to the votes of 10 configurations, mostly of earlier model versions, on a calibration corpus of 100 records, 21 relevant (Appendix A).

### 2.3 Data sets and reference standards

The derivation set came from SYNERGY, an open data set of systematic reviews with record-level decisions [27]. Of 14 health reviews with recoverable criteria, one was excluded for an incoherent reference standard under an amendment prompted by its own screening data (Appendix C). From each of the remaining 13 reviews we took all relevant records up to 45 and an equal number of irrelevant ones at random (724 records, 364 relevant). The reference standard was final inclusion.

Both replications used Cochrane reviews from the Conference and Labs of the Evaluation Forum (CLEF) eHealth Technology Assisted Reviews (TAR) 2019 collection, whose records carry a title–abstract label and a final-inclusion label [28]. Replication 1 took five diagnostic test accuracy (DTA) and five intervention reviews in a registered pseudo-random order (740 records, 370 relevant); replication 2 took 16 new reviews (626 records, 313 relevant). The reference standard was the title–abstract label.

At real prevalence, we screened 11 complete CLEF collections without sampling (5,919 records; 217 relevant at title–abstract level and 96 finally included) and the rebuilt PubMed searches of six reviews published in July–October 2026, after the documented training cut-off of Claude Haiku 5.5 [29] (6,277 records; 178 of 208 included-study records, as 30 lay outside the rebuilt searches). The 2026 reviews were chosen by criteria fixed before searching, among openly licensed reviews (Appendix B). Sealed decision rules used final inclusion, reading load and the title–abstract label, which the CLEF seals named as the primary reference.

Eligibility criteria were transcribed in Spanish into a fixed template, blind to labels and votes, by LLM agents (Claude, Anthropic) under the authors’ supervision: from the full-text methods section in eight of the 13 derivation reviews and from the published abstract in all others.

### 2.4 Models and calling route

Eleven models from four developers (Table 2) voted in the derivation study and in replication 1. In these two stages the Anthropic models voted as isolated agents in Claude Code and the others through the OpenRouter application programming interface (API); from replication 2 onwards every model was called through an API. To test the calling route, Haiku and Sonnet re-voted the derivation batches, and Haiku the replication 1 batches, through the Anthropic API (not pre-specified). Records were sent in English, in batches of up to 50, with the same system prompt, criteria, output schema and sensitivity instruction (missing or vague information counts as met when the design is eligible; settings in Appendix B). Every batch was checked for completeness and resubmitted once if incomplete; a decision still missing counted as “include”.

**Table 2.** Sensitivity and false-positive rate of each model voting alone.

| **Model** | **Derivation (364 relevant): sensitivity (95% CI)** | **FPR** | **Replication 1 (370 relevant): sensitivity (95% CI)** | **FPR** | **Replication 2, API (313 relevant): sensitivity (95% CI)** | **FPR** | **US\$ per 1,000 records** |
|---|---|---|---|---|---|---|---|
| Claude Haiku 5.5 | 0.887 (0.851–0.916) | 0.218 | 0.870 (0.832–0.901) | 0.167 | 0.821 (0.775–0.860) | 0.083 | 0.12–0.14 |
| Claude Sonnet 5.5 | 0.871 (0.833–0.901) | 0.090 | 0.873 (0.835–0.903) | 0.091 | 0.741 (0.690–0.787) | 0.029 | 1.95–2.83 |
| Claude Opus 5.5 | 0.868 (0.829–0.899) | 0.107 | 0.865 (0.826–0.896) | 0.105 | — | — | 5.55–5.66 |
| Claude Fable 5.1 | 0.920 (0.888–0.944) | 0.146 | 0.873 (0.835–0.903) | 0.148 | — | — | 13.86–14.14 |
| Gemini 3.5 Flash-Lite | 0.885 (0.848–0.913) | 0.334 | 0.919 (0.887–0.943) | 0.492 | 0.927 (0.892–0.951) | 0.317 | 0.58–0.59 |
| Gemini 3.8 Flash | 0.819 (0.776–0.855) | 0.103 | 0.862 (0.823–0.894) | 0.144 | — | — | 1.00–1.02 |
| Gemini 4 Argon | 0.857 (0.817–0.889) | 0.114 | 0.862 (0.823–0.894) | 0.122 | — | — | 2.65–2.72 |
| GPT-6 Luna | 0.690 (0.640–0.735) | 0.153 | 0.832 (0.791–0.867) | 0.161 | 0.633 (0.578–0.684) | 0.110 | 0.13 |
| GPT-6 Sol | 0.810 (0.767–0.847) | 0.142 | 0.827 (0.785–0.862) | 0.116 | — | — | 2.49–2.52 |
| DeepSeek V4.1 Flash | 0.769 (0.723–0.810) | 0.131 | 0.873 (0.835–0.903) | 0.125 | 0.799 (0.751–0.839) | 0.102 | 0.22–0.26 |
| DeepSeek V4 Pro | 0.863 (0.823–0.894) | 0.257 | 0.878 (0.841–0.908) | 0.407 | 0.840 (0.796–0.877) | 0.293 | 0.74–0.77 |

CI, Wilson confidence interval; FPR, false-positive rate among irrelevant records, reweighted to each review's screened corpus (derivation) or full collection (replications); API, application programming interface. Reference standard: final inclusion (derivation) and title–abstract label (replications). Derivation values for Claude Haiku 5.5 and Sonnet 5.5 are their first agent runs, on batches of the full sampled corpus (Appendix B.1); Section 3.4 compares their second agent runs, on the paired batches, with the API. In the derivation set and replication 1, the Anthropic models voted as agents, and their cost is the list price applied to tokens measured for the other models (probably an underestimate); in replication 2, Anthropic costs are batch list prices applied to logged tokens; all other costs are the amounts billed by OpenRouter. Costs are ranges across the stages.

### 2.5 Aggregation rules and the evaluated pipeline

We evaluated every subset of one to five models (six in replication 2) under “any model includes” and, for odd sizes, under sequential majority, in which models are called until one side has a majority; for three models, two vote and the third adjudicates their disagreements. The pipeline evaluated at real prevalence was pre-specified on 28 October 2026, before any vote of replication 2: Claude Haiku 5.5 and DeepSeek V4.1 Flash vote on every record, and DeepSeek V4 Pro adjudicates disagreements and records with one missing vote. We compared it with Haiku alone and with “either voter includes”.

### 2.6 Outcomes, cost and optimal size

Sensitivity was the proportion of relevant records sent to human reading. The false-positive rate (FPR) was computed over irrelevant records reweighted to the screened corpus (derivation) or full collection (replications) of each review. At real prevalence we report the reading load, the proportion of all records sent to a human. Machine cost per 1,000 records was the amount billed by OpenRouter or, for Anthropic models, list price applied to logged tokens (for agents, to tokens measured for other models); one human reading was costed at 38 seconds at US$25 per hour (US$0.264).

Net benefit per record [30], in human readings, was NB = π · sensitivity · V/h − (1 − π) · FPR − machine cost/h, where π is prevalence, V the value of finding a relevant study and h the cost of one reading. We used π of 2%, 5% and 10% and V/h of 100–1,000 readings; double human screening implies 180–1,600 (Appendix A). In each bootstrap replicate (reviews, then records within reviews; 200 replicates, 1,000 in replication 2) we recorded K95, the smallest number of models whose best configuration reached 95% of the maximum gain over the best single model.

### 2.7 Statistical analysis

Proportions have Wilson 95% confidence intervals (CI); paired comparisons used exact McNemar tests. Agreement between models was measured with Fleiss’ kappa, separately on relevant and irrelevant records (self-agreement: Cohen’s kappa over all records), and the within-versus between-developer difference with a two-stage bootstrap CI. Each prediction was judged by its sealed rule: supported, not supported or, where the rule provided, indeterminate. Totals pooled across the 17 reviews and the latent class analysis were post hoc.

## 3. Results

### 3.1 Theory and calibration

The fitted model gave a mean single-model sensitivity of 0.885 and an error correlation on relevant records of ρ = 0.62. Under “any model includes” it predicted 11.5 missed relevant records per 100 with one model, 7.6 with two, 6.0 with three and 5.1 with four, against 1.3 and 0.2 with two and three independent models (Fig. 2A). It underestimated observed misses with one to three models (observed mean single-model sensitivity 0.862), matched them at four and overestimated them beyond. With ρ = 0.62, no ensemble of exchangeable models is worth more than 1.6 independent ones (Fig. 2B), and a majority of an odd number of models cannot exceed a sensitivity of 0.899.

**Fig. 2.**
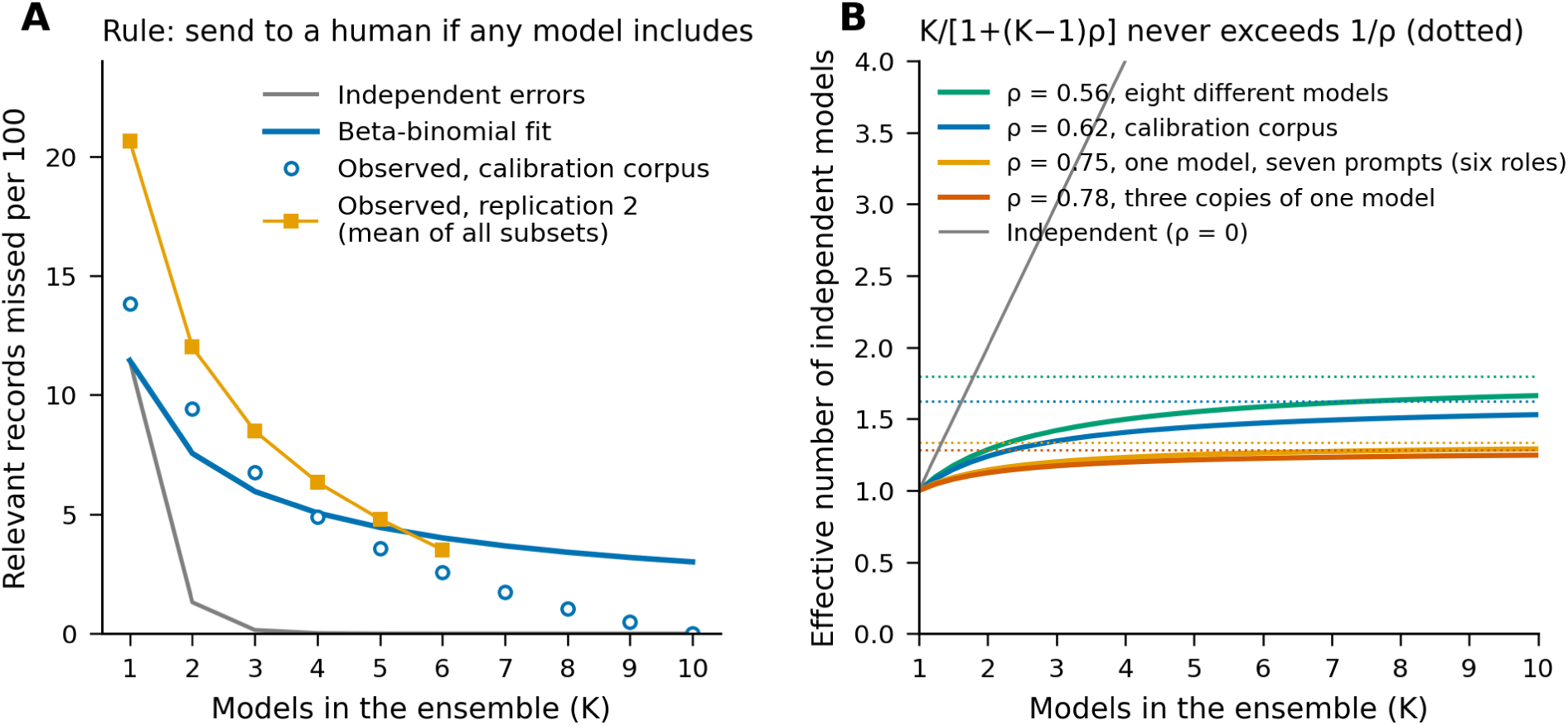
Why ensembles saturate. (A) Relevant records missed per 100 under “any model includes”, as a function of the number of models: independent errors, the beta-binomial model fitted to the calibration corpus (mean single-model sensitivity 0.885, ρ = 0.62), the observed mean over all subsets in the calibration corpus, and the observed mean over all subsets in replication 2. (B) Effective number of independent models, K/[1 + (K − 1)ρ], for the correlations observed among eight different models, in the calibration corpus, among seven prompts of one model (standard and six roles) and among three copies of one model; dotted lines show the ceiling 1/ρ.

### 3.2 Derivation

Single-model sensitivity ranged from 0.690 to 0.920 (Table 2). Votes on relevant records agreed more within than between developers (mean kappa 0.538 vs 0.414; difference 95% CI 0.01–0.25), but the excess came from the Anthropic pairs, which also shared the agent route. Three or fewer models reached 95% of the attainable gain in at least 86% of replicates in every scenario (Fig. 3). Under “any model includes”, the best ensemble of each size, selected in the same data, reached 0.920 with one model, 0.975 with two and 0.981 with three, four and five models (Fig. 4A), while the FPR rose from 0.146 to 0.459. The best pair, Claude Haiku 5.5 and Gemini 3.5 Flash-Lite, found 19 relevant records that the Haiku– Sonnet pair missed and missed one it found (P < 0.001), with almost twice its FPR (0.418 vs 0.223).

**Fig. 3.**
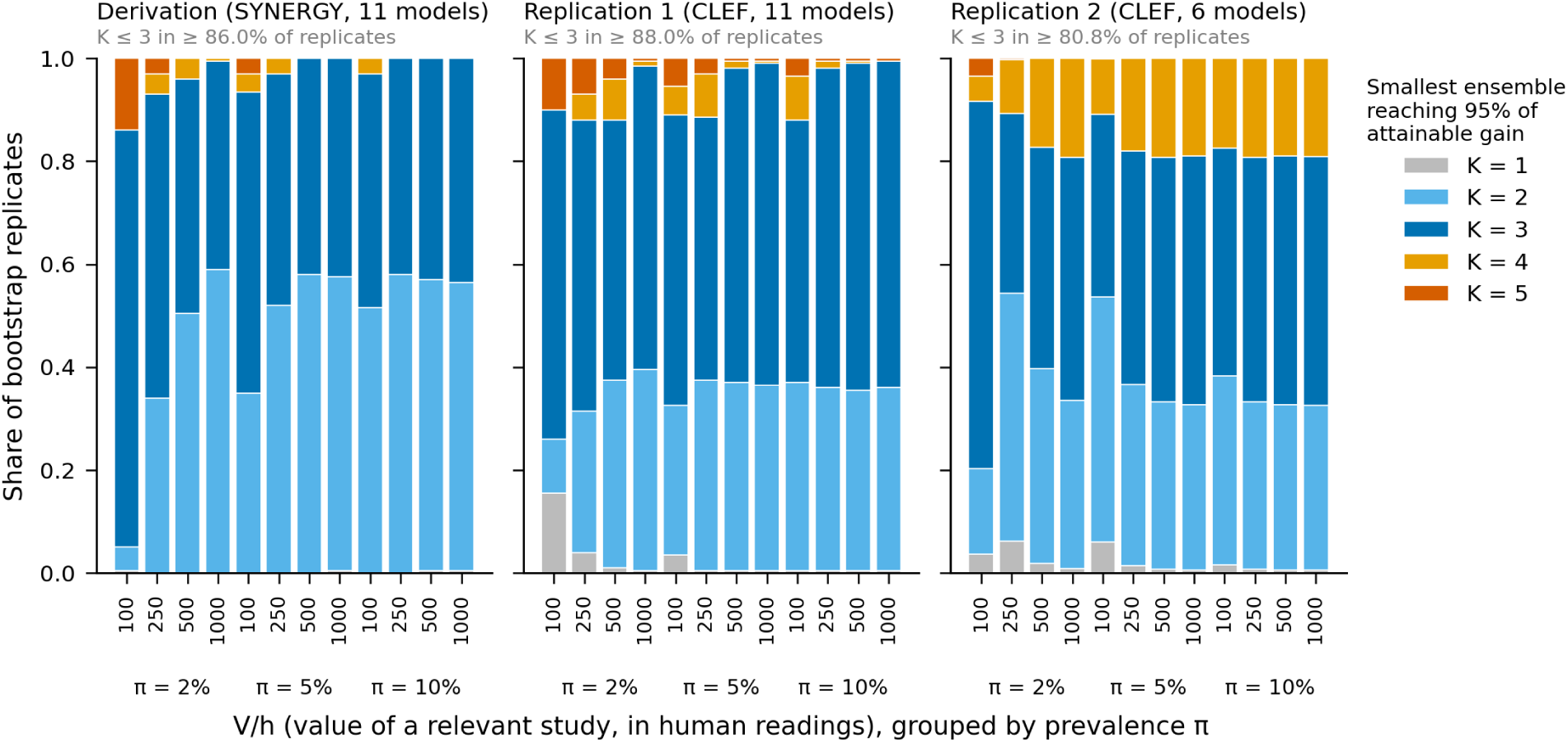
Smallest ensemble reaching 95% of the attainable gain in net benefit over the best single model (K95), by scenario. Bars show the share of bootstrap replicates (200 in the derivation and replication 1; 1,000 in replication 2) in which each K was selected, for each prevalence (π) and value of a relevant study in human readings (V/h). Candidates were all subsets of one to five models (one to six in replication 2) under “any model includes” and, for odd sizes, sequential majority; the best configuration of each size was chosen within each replicate.

**Fig. 4.**
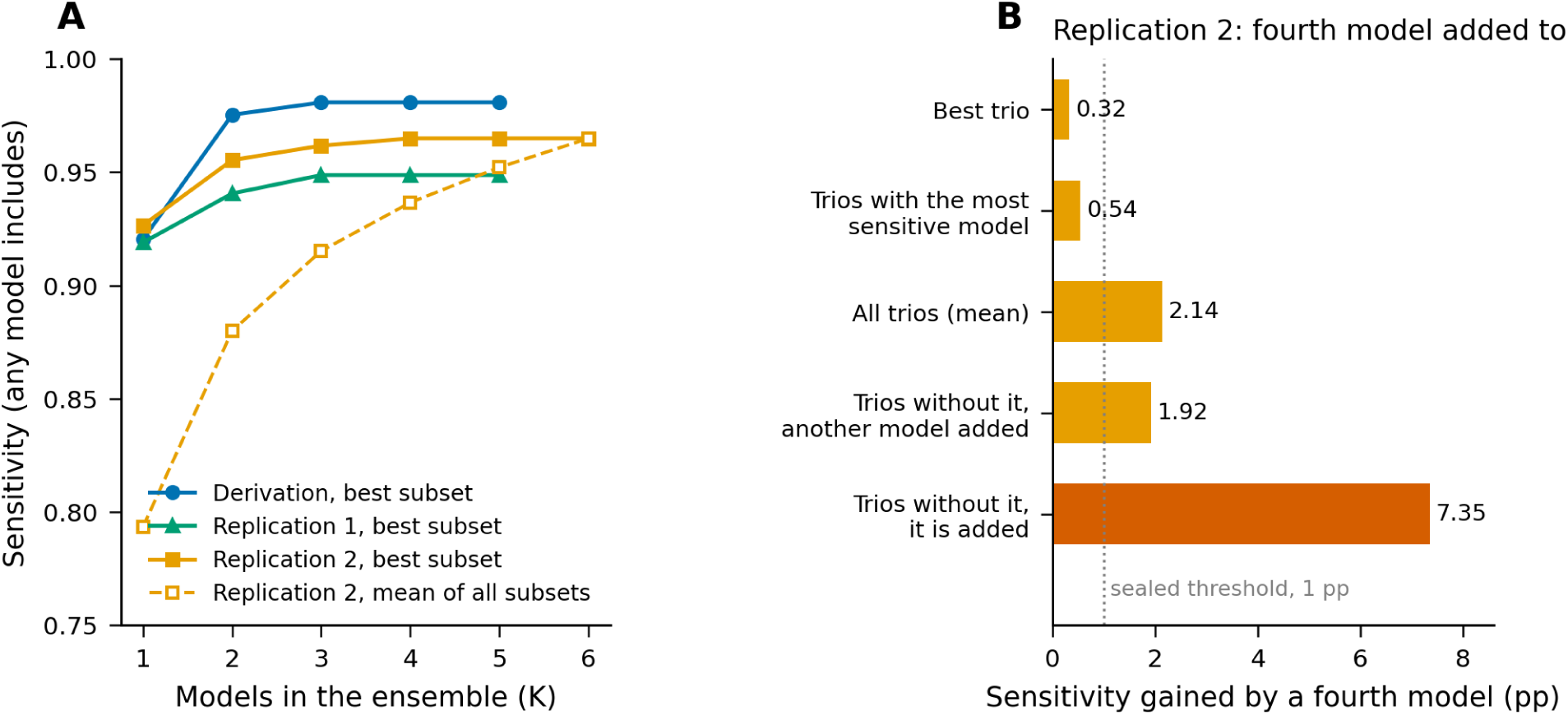
Gain from adding models. (A) Sensitivity under “any model includes” of the best subset of each size in the three data sets, and of the mean over all subsets in replication 2. (B) Replication 2: sensitivity gained by adding a fourth model to the best trio, to trios that already contained the most sensitive model (Gemini 3.5 Flash-Lite), to all trios on average, and to trios without it when another model or Flash-Lite itself was added; the dotted line marks the sealed threshold of 1 percentage point (pp).

### 3.3 Replication 1

Six of seven registered predictions were supported (Table 3); the size prediction failed in one of 12 scenarios (π = 2%, V/h = 100), where a single model sufficed in 15.5% of replicates.

**Table 3.** Predictions fixed before each data set was opened, and results.

| Stage | Prediction (criterion) | Result | Supported |
|---|---|---|---|
| Replication 1 (registered) | P1 Two or three models in ≥ 80% of replicates in every scenario | 84%–99% in 11 scenarios; 74.5% at π = 2%, V/h = 100 | No |
|  | P2 Four or more models in ≤ 15% of replicates in every scenario | Maximum 12% | Yes |
|  | P3 Within-developer kappa > between-developer kappa (CI excludes 0) | 0.756 vs 0.710 (95% CI of difference 0.008–0.094) | Yes |
|  | P4 Best pair under "any model includes" spans two developers | DeepSeek V4.1 Flash + Gemini 3.5 Flash-Lite | Yes |
|  | P5 Inexpensive adjudicator within ±3 pp of Fable 5.1, at lower cost | 0.889 vs 0.878 (DeepSeek V4 Pro and Flash-Lite tied) | Yes |
|  | P6 Fourth model adds < 1 pp to the best trio | 0.0 pp | Yes |
|  | P7 Each repeated model agrees more with itself than with others | 6 of 6 models | Yes |
| Replication 2 (sealed) | R1 Three or fewer models in ≥ 80% of replicates in every scenario | Minimum 80.8%; reversed with about a third of other bootstrap seeds | Yes, narrowly |
|  | R2 Fourth model adds < 1 pp to trios on average | 2.14 pp (95% CI 1.24–3.07); by the sealed rule, the size recommendation then depends on the calling route | No |
|  | R3 Inexpensive adjudicators within ±2 pp of DeepSeek V4 Pro | Flash-Lite +1.3 pp; DeepSeek V4.1 Flash −2.6 pp | No |
|  | R4 Haiku–Sonnet kappa > between-developer kappa (CI excludes 0) | 0.692 vs 0.426 (95% CI of difference 0.086–0.457) | Yes |
|  | R5 Rank of six configurations kept (Spearman r<sub>s</sub> ≥ 0.80) | r<sub>s</sub> = 0.87 | Yes |
| CLEF collections, batch 1 (sealed) | V1 ≥ 80% of title–abstract relevant records reach reading | 42/65 (64.6%) | No |
|  | V2 ≥ 90% of included studies reach reading | 21/22 (95.5%) | Yes |
|  | V3 Reading load ≤ 20% | 7.2% | Yes |
| CLEF collections, batch 2 (sealed) | W1 ≥ 90% of included studies reach reading | 73/74 (98.6%) | Yes |
|  | W2 Reading load ≤ 20% | 13.1% | Yes |
|  | W3 ≥ 70% of title–abstract relevant records reach reading | 131/152 (86.2%) | Yes |
| Batch composition, CLEF batch 2 (sealed) | X1–X2 Balanced batches send more records to reading than real-prevalence batches | 0.0 pp in relevant and irrelevant records | No |
| Reviews of 2026 (sealed) | Z1 ≥ 90% of included studies reach reading | 163/178 (91.6%) | Yes |
|  | Z2 Reading load ≤ 20% | 12.6% | Yes |
|  | Z3 No more than 5 pp below the CLEF collections | −6.3 pp (90% interval −13.1 to +0.6) | No (sealed reading indeterminate) |

pp, percentage points; CI, confidence interval; π, prevalence; V/h, value of a relevant study in human readings. W1–W3 were written after the batch 1 results were known. Comparisons with the best trio (Fig. 4B) are post hoc. X1–X2 tested whether the composition of batches changes decisions (same records screened in balanced and in real-prevalence batches); the test had low power.

### 3.4 Calling route and replication 2

On identical derivation batches, Claude Haiku 5.5 called through the API was 3.8 percentage points (pp) less sensitive than in a second agent run (0.871 to 0.832; P = 0.049) and Sonnet 6.0 pp less (0.879 to 0.819; P = 0.001); the route also changed effort and output settings. On identical CLEF batches, Haiku barely changed (0.870 vs 0.862; P = 0.69).

In replication 2, with all models called through an API, three of five sealed predictions were supported (Table 3). Agreement remained higher within developers (Haiku–Sonnet kappa 0.69 vs 0.43), though Haiku–DeepSeek V4.1 Flash reached 0.72. Three or fewer models reached 95% of the attainable gain in at least 80.8% of replicates in every scenario (Fig. 3), just above the 80% threshold; with about a third of other bootstrap seeds this verdict reversed. A fourth model added 2.14 pp (95% CI 1.24–3.07) to trios on average, above the sealed threshold of 1 pp, but 0.32 pp to the best trio, which already contained the most sensitive model, Gemini 3.5 Flash-Lite (Fig. 4). As inexpensive adjudicators, Flash-Lite matched DeepSeek V4 Pro (+1.3 pp) but DeepSeek V4.1 Flash did not (−2.6 pp). Through the API, Sonnet was the most specific model (sensitivity 0.741; FPR 0.029), and Haiku plus Sonnet with DeepSeek V4 Pro adjudicating matched Haiku alone (0.815 vs 0.821) at 18 times the cost.

### 3.5 The pipeline at real prevalence

In the 11 complete CLEF collections (Table 4; Fig. 5), the pipeline sent 94 of 96 included studies (97.9%; 95% CI 92.7–99.4) and 173 of 217 records relevant at title–abstract level (79.7%) to human reading, with a reading load of 11.6%. Sealed thresholds for included studies and reading load were met; for title– abstract relevant records, an 80% floor failed in the first batch and a 70% floor, set after it, held in the second (Table 3). In the six reviews published in 2026, the pipeline sent 163 of 178 included studies (91.6%; 95% CI 86.6–94.8) with a load of 12.6%, meeting the pre-specified thresholds (≥ 90% and ≤ 20%); the proportion was 6.3 pp lower than in the CLEF collections, so non-inferiority within 5 pp was not shown. All 36 included records of the Cochrane review were found, against 127 of 142 in the five journal reviews. In a post hoc, unblinded classification by an LLM reader, two of the 15 misses were clear model errors, one was debatable, 11 involved eligibility rules not stated in the review’s abstract and one was a reference-standard error; three had been included by Haiku and excluded by the adjudicator (Appendix D).

**Table 4.** The pipeline and its components at real prevalence.

| Configuration | 11 CLEF collections: included reaching reading | Title–abstract relevant reaching reading | Reading load | US\$ per 1,000 records | 6 reviews of 2026: included reaching reading | Reading load | US\$ per 1,000 records |
|---|---|---|---|---|---|---|---|
| Pipeline: Haiku + DeepSeek V4.1 Flash, DeepSeek V4 Pro adjudicates | 94/96 (97.9%) | 173/217 (79.7%) | 11.6% | 0.51 | 163/178 (91.6%) | 12.6% | 0.48 |
| Claude Haiku 5.5 alone | 94/96 (97.9%) | 165/217 (76.0%) | 10.7% | 0.11 | 165/178 (92.7%) | 13.2% | 0.11 |
| DeepSeek V4.1 Flash alone | 94/96 (97.9%) | 163/217 (75.1%) | 9.9% | 0.36 | 162/178 (91.0%) | 12.2% | 0.31 |
| Either voter includes | 94/96 (97.9%) | 173/217 (79.7%) | 13.5% | 0.46 | 166/178 (93.3%) | 15.6% | 0.43 |
| Both voters include | 94/96 (97.9%) | 155/217 (71.4%) | 7.1% | 0.46 | 161/178 (90.4%) | 9.8% | 0.43 |

Reading load: proportion of all records sent to a human reader (5,919 records in CLEF; 6,277 in 2026). For single voters and the "either" and "both" rules, a missing vote counted as "include"; in the pipeline, records with a missing vote went to the adjudicator. OpenRouter costs as billed; Claude Haiku 5.5 at Anthropic batch list price applied to logged tokens.

**Fig. 5.**
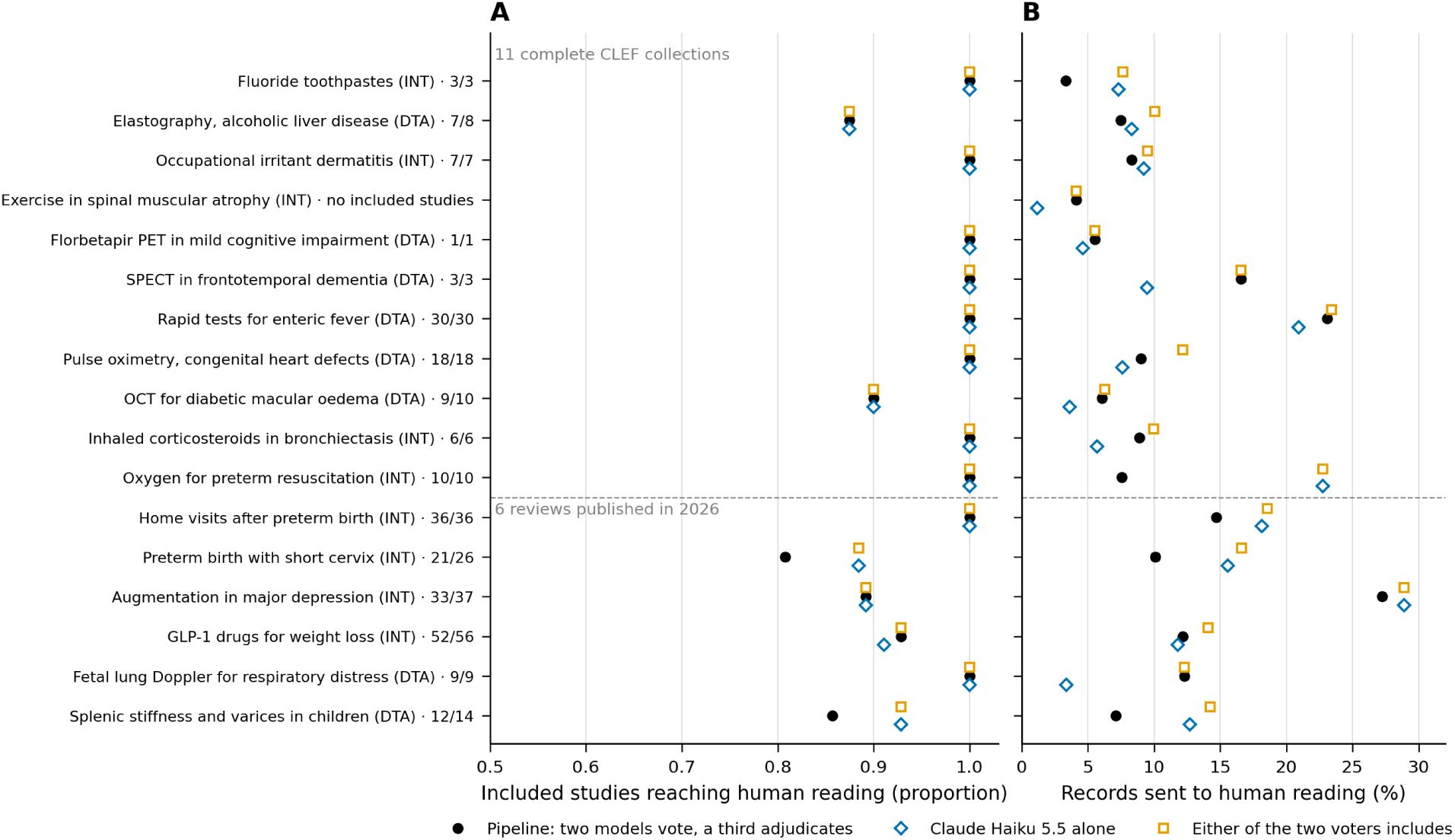
The pipeline at real prevalence, by review. (A) Proportion of included studies that reached human reading, with the count for the pipeline. (B) Proportion of all records sent to human reading. Pipeline: Claude Haiku 5.5 and DeepSeek V4.1 Flash vote; DeepSeek V4 Pro adjudicates disagreements and records with one missing vote (in the fetal lung Doppler review, a failed batch raised the pipeline’s load; Appendix D). Included: in the 2026 reviews, PubMed records of included studies. DTA, diagnostic test accuracy; INT, intervention; PET, positron emission tomography; SPECT, single-photon emission computed tomography; OCT, optical coherence tomography; GLP-1, glucagon-like peptide-1. One CLEF collection had no included study.

Across the 17 reviews, the pipeline sent 257 of 274 included studies (93.8%) to human reading, with 12.1% of 12,196 records sent. Claude Haiku 5.5 alone sent 259 (94.5%) with 12.0%, and “either voter includes” 260 (94.9%) with 14.6%; compared with the latter, adjudication saved 301 readings at the cost of three included studies, about 100 readings per study. Haiku alone cost US$0.11 per 1,000 records, 21%–24% of the pipeline’s cost (US$0.48–0.51), whereas human reading at a 12% load costs about US$32.

### 3.6 Further analyses

Seven Claude Haiku 5.5 API calls (standard prompt plus six role prompts) behaved like 1.3 independent models (ρ = 0.75; three copies, ρ = 0.78; eight different models, ρ = 0.56). Going from three to seven raised “any” sensitivity by 2.1 pp (90% CI 1.2–3.6) and the FPR by 3.3 pp, paying for the extra false positives in 7 of 8 sealed scenarios (sealed overall reading indeterminate); majority voting did not gain (Appendix E). Giving a model the methods-section criteria recovered four of eight targeted missed studies (one of eight on a same-day re-vote with abstract criteria), below the pre-specified six.

## 4. Discussion

### 4.1 Principal findings

Correlation, not the number of models, set the useful size of a screening ensemble: eight different models carried the information of about 1.6 independent ones, and the best pair or trio captured 95% of the attainable gain in most replicates of three data sets. Which models were combined and how mattered as much: a fourth model added little to a good trio and more to a poor one, inexpensive adjudicators differed, and in one data set sensitivity was lower through an API. At real prevalence, the pipeline’s more sensitive voter alone sent as many included studies to reading at under a quarter of the machine cost.

### 4.2 Comparison with previous studies

These results agree with the theory of correlated juries [18–20] and with the finding that nine LLM judges were worth about two independent votes [22]. Under “any model includes”, ensembles gained sensitivity at the expense of specificity [9,11,12,14]; agreement rules gained accuracy but left records undecided [10,13]. We add an explicit optimisation, tested with predictions fixed in advance. Size predictions held in part: four or more models were rarely selected, but one model sufficed in some replicates and a fourth model still improved average trios; adjudicator predictions held in replication 1 but not in replication 2. Agreement was higher within developers, but a cross-developer pair could be as correlated, and mixing developers was not shown to help against a post hoc latent reference (Appendix E).

### 4.3 Implications

Four practical rules follow. First, in our data the best pair or trio captured most of the attainable gain; more models helped mainly poorly chosen trios. A third model can adjudicate disagreements, reproducing a majority of three with fewer calls, but at real prevalence this saved about 100 readings per included study lost (three studies; post hoc), below the value implied by double screening, so letting either voter include may be preferable. Second, models should be chosen by measuring their sensitivity and error correlation on a local labelled set, through the route and settings to be used in production, rather than by size, price or developer. Third, every ensemble should be compared with its best single member on sensitivity and reading load; in our evaluation, the pipeline sent no more included studies to reading than Haiku alone. Fourth, output completeness must be checked, because models dropped records or returned empty outputs, and a missing vote must never count as an exclusion. These rules fit current guidance that artificial intelligence in evidence synthesis be justified, validated and reported [31].

Model choice is not the only limit: methods-section criteria did not clearly recover the misses attributed post hoc to unstated rules, and three were lost because the adjudicator overruled an inclusion.

### 4.4 Strengths and limitations

Strengths are the paired design, 11 models from four developers, predictions fixed before each new data set was opened, and an evaluation at real prevalence including reviews published after the main voter’s training cut-off.

The reference standard is the published review, whose errors count against all models at once, inflating correlation; blinded human adjudication of the pipeline’s misses is planned as a separate study. Only the first replication was registered publicly; later stages were sealed with local hashes and no external time-stamp, and the derivation analysis was exploratory. K95 used each sample’s best configuration; a trio chosen on other reviews fell 2.3–3.4 pp short of eight models (Appendix E). LLM agents transcribed the eligibility criteria and, for the 2026 reviews, extracted the included-study lists. Variation between reviews is the main source of uncertainty, samples were half relevant, and each analysis used one run per model. SYNERGY and CLEF reviews predate all models, and two DeepSeek models recalled some included CLEF studies from memory (Appendix E); the lower yield in the 2026 reviews may reflect review source rather than date, because all misses were in journal reviews. Batch size can affect LLM screening [32]. Model versions change quickly, so accuracies must be re-measured. The authors develop the evaluated pipeline.

### 4.5 Conclusion

Because LLM errors are correlated, our results suggest that two or three well-chosen models may suffice for title–abstract screening, but this needs deeper exploration in future studies. Which models are chosen and how their votes are combined matter as much as how many, and any ensemble should be compared with its best single member.

## CRediT authorship contribution statement

**Eduardo Quiñelen:** Conceptualization, Methodology, Software, Formal analysis, Investigation, Data curation, Validation, Visualization, Resources, Writing – original draft, Writing – review and editing. **Ignacio Neumann:** Methodology, Supervision, Writing – review and editing. **Miguel Contreras:** Conceptualization, Software, Resources.

## Ethics statement

Ethics approval was not required because the study used published bibliographic records and review decisions and involved no human participants.

## Data availability

SYNERGY (CC0 1.0) [27] and CLEF TAR 2019 (MIT licence) [28,33] are publicly available. Record identifiers (PubMed or OpenAlex) with reference labels, transcribed eligibility protocols, every model vote of the reported analyses, batch compositions, prompts, cost logs, sealed prediction files with their hashes and the analysis code, with a script that recomputes the main results, are deposited in Zenodo (https://doi.org/10.5281/zenodo.24543734). Titles and abstracts of screened records are not redistributed, except the citations of the included studies of the 2026 reviews; the calibration corpus, from an unpublished review, and internal working documents of the role-agent stage are not deposited, the latter represented by their hashes. Votes and labels of the sealed third replication will be added when analysed.

## Declaration of generative AI and AI-assisted technologies in the manuscript preparation process

Large language models were the object of study (Section 2.4) and assisted in study procedures: they transcribed eligibility criteria (Section 2.3), extracted the included-study lists of the 2026 reviews and classified missed studies post hoc (Appendices B and D). During the preparation of this work the authors also used Claude (Anthropic; Claude Opus 5.5 in Claude Code) to write analysis and figure code, to check the numbers in the manuscript against the source files and to draft the English text. After using this tool, the authors reviewed and edited the content as needed and take full responsibility for the content of the published article.

## Acknowledgements

None.

## References

[1] Lefebvre C, Glanville J, Briscoe S, Featherstone R, Littlewood A, Metzendorf MI, et al. Chapter 4: searching for and selecting studies [last updated March 2025]. In: Higgins JPT, Thomas J, Chandler J, Cumpston M, Li T, Page MJ, et al., editors. Cochrane handbook for systematic reviews of interventions, version 6.5.1. Cochrane; 2025. https://www.cochrane.org/authors/handbooks-and-manuals/handbook/current/chapter-04 [accessed 10 November 2026].

[2] Gartlehner G, Affengruber L, Titscher V, Noel-Storr A, Dooley G, Ballarini N, et al. Single-reviewer abstract screening missed 13 percent of relevant studies: a crowd-based, randomized controlled trial. J Clin Epidemiol 2020;121:20–8. 10.1016/j.jclinepi.2020.01.005.

[3] Waffenschmidt S, Knelangen M, Sieben W, Bühn S, Pieper D. Single screening versus conventional double screening for study selection in systematic reviews: a methodological systematic review. BMC Med Res Methodol 2019;19:132. 10.1186/s12874-019-0782-0.

[4] Stoll CRT, Izadi S, Fowler S, Green P, Suls J, Colditz GA. The value of a second reviewer for study selection in systematic reviews. Res Synth Methods 2019;10:539–45. 10.1002/jrsm.1369.

[5] Lieberum JL, Toews M, Metzendorf MI, Heilmeyer F, Siemens W, Haverkamp C, et al. Large language models for conducting systematic reviews: on the rise, but not yet ready for use—a scoping review. J Clin Epidemiol 2025;181:111746. 10.1016/j.jclinepi.2025.111746.

[6] Laignelot F, Martin GL, Ossman M, Pingeon O, Boubaker A, Picovschi E, et al. Large language models show promising performance for some systematic review tasks but call for cautious implementation: a systematic review. J Clin Epidemiol 2026;194:112221. 10.1016/j.jclinepi.2026.112221.

[7] Xie C, Kong W, Pi L, Qi D, Yang Y, Wang B, et al. Performance of large language models in automated medical literature screening: a systematic review and meta-analysis. J Evid Based Med 2026;19:e70166. 10.1111/jebm.70166.

[8] Cao C, Sang J, Arora R, Chen D, Kloosterman R, Cecere M, et al. Development of prompt templates for large language model-driven screening in systematic reviews. Ann Intern Med 2025;178:389–401. 10.7326/ANNALS-24-02189.

[9] Sanghera R, Thirunavukarasu AJ, El Khoury M, O’Logbon J, Chen Y, Watt A, et al. High-performance automated abstract screening with large language model ensembles. J Am Med Inform Assoc 2025;32:893–904. 10.1093/jamia/ocaf050.

[10] Courvoisier DS, Buitrago-Garcia D, Buclin CP, Bürgisser N, Iudici M, Mongin D. Beyond human gold standards: a multimodel framework for automated abstract classification and information extraction. Res Synth Methods 2026;17:365–77. 10.1017/rsm.2025.10054.

[11] Fagerberg P, Sallander O, Vikhe Patil K, Thunborg C, Lundström L, Berg A, et al. Audited large language model triage for systematic review screening in national clinical guideline production: validation and prospective deployment [preprint]. medRxiv 2026. 10.64898/2026.06.02.26354724.

[12] Parmar M, Naqvi SAA, Warraich K, Saeidi A, Rawal S, Faisal KS, et al. Collaborative large language models (LLMs) are all you need for screening in systematic reviews [preprint]. medRxiv 2026. 10.64898/2026.02.07.26345640.

[13] Hilkenmeier F, Stoltenberg M, Stierle C. Using full agreement across multiple large language models for title-and-abstract screening in systematic reviews: a proof-of-concept. Syst Rev 2026;15:191. 10.1186/s13643-026-03228-4.

[14] Oami T, Okada Y, Nakada TA. Optimal large language models to screen citations for systematic reviews. Res Synth Methods 2025;16:859–75. 10.1017/rsm.2025.10014.

[15] Boland PJ. Majority systems and the Condorcet jury theorem. Statistician 1989;38:181–9. 10.2307/2348873.

[16] Kim EM, Garg A, Peng K, Garg N. Correlated errors in large language models. In: Proceedings of the 42nd International Conference on Machine Learning; 2025 Jul 13-19; Vancouver, Canada. Proc Mach Learn Res 2025;267:30038–66. https://proceedings.mlr.press/v267/kim25e.html.

[17] Goel S, Strüber J, Auzina IA, Chandra KK, Kumaraguru P, Kiela D, et al. Great models think alike and this undermines AI oversight. In: Proceedings of the 42nd International Conference on Machine Learning; 2025 Jul 13-19; Vancouver, Canada. Proc Mach Learn Res 2025;267:19621–78. https://proceedings.mlr.press/v267/goel25b.html.

[18] Ladha KK. The Condorcet jury theorem, free speech, and correlated votes. Am J Pol Sci 1992;36:617–34. 10.2307/2111584.

[19] Kaniovski S. Aggregation of correlated votes and Condorcet’s jury theorem. Theory Decis 2010;69:453–68. 10.1007/s11238-008-9120-4.

[20] Kaniovski S, Zaigraev A. Optimal jury design for homogeneous juries with correlated votes. Theory Decis 2011;71:439–59. 10.1007/s11238-009-9170-2.

[21] Gardner IA, Stryhn H, Lind P, Collins MT. Conditional dependence between tests affects the diagnosis and surveillance of animal diseases. Prev Vet Med 2000;45:107–22. 10.1016/S0167-5877(00)00119-7.

[22] Kohli G. Nine judges, two effective votes: correlated errors undermine LLM evaluation panels [preprint]. arXiv 2026. 10.48550/arXiv.2605.29800.

[23] Li H, Hai M. State-dependent error correlations shape voting thresholds in committees of AI agents [preprint]. arXiv 2026. 10.48550/arXiv.2607.23931.

[24] Quiñelen E. Optimal size of LLM ensembles for systematic review [registration]. OSF Registries; 2026. https://osf.io/zakg9.

[25] Bossuyt PM, Reitsma JB, Bruns DE, Gatsonis CA, Glasziou PP, Irwig L, et al. STARD 2015: an updated list of essential items for reporting diagnostic accuracy studies. BMJ 2015;351:h5527. 10.1136/bmj.h5527.

[26] Gallifant J, Afshar M, Ameen S, Aphinyanaphongs Y, Chen S, Cacciamani G, et al. The TRIPOD-LLM reporting guideline for studies using large language models. Nat Med 2025;31:60–9. 10.1038/s41591-024-03425-5.

[27] De Bruin J, Ma Y, Ferdinands G, Teijema J, Van de Schoot R. SYNERGY – open machine learning dataset on study selection in systematic reviews. Version 1.0 [dataset]. DataverseNL; 2023. 10.34894/HE6NAQ.

[28] Kanoulas E, Li D, Azzopardi L, Spijker R. CLEF 2019 technology assisted reviews in empirical medicine overview. In: Cappellato L, Ferro N, Losada DE, Müller H, editors. Working Notes of CLEF 2019 – Conference and Labs of the Evaluation Forum; 2019 Sep 9-12; Lugano, Switzerland. CEUR Workshop Proceedings, vol. 2380; 2019. https://ceur-ws.org/Vol-2380/paper_250.pdf.

[29] Anthropic. Models overview, https://platform.claude.com/docs/en/models/overview; 2026 [accessed 10 November 2026].

[30] Vickers AJ, Elkin EB. Decision curve analysis: a novel method for evaluating prediction models. Med Decis Making 2006;26:565–74. 10.1177/0272989X06295361.

[31] Flemyng E, Noel-Storr A, Macura B, Gartlehner G, Thomas J, Meerpohl JJ, et al. Position statement on artificial intelligence (AI) use in evidence synthesis across Cochrane, the Campbell Collaboration, JBI, and the Collaboration for Environmental Evidence 2025. JBI Evid Synth 2025;23:2162–6. 10.11124/JBIES-25-00480.

[32] Fagerberg P, Sallander O, Vikhe Patil K, Berg A, Nyman A, Borg N, et al. Batch size effects on mid-2025 state-of-the-art large language model performance in automated title and abstract screening. Cochrane Evid Synth Methods 2026;4:e70082. 10.1002/cesm.70082.

[33] CLEF-TAR. Technologically assisted reviews in empirical medicine [dataset]. GitHub; 2019. https://github.com/CLEF-TAR/tar [accessed 10 November 2026].

