## Supplementary appendix (Appendices A-G) for "How many large language models does title–abstract screening need? Theory, two replications and an evaluation at real prevalence"

Appendices A–G. Unprefixed section, table, figure and reference numbers (e.g., Section 2.3, Table 2, Fig. 1, [26]) refer to the main text; references numbered S1–S3 are listed at the end of this supplement.

### Appendix A. Theory, calibration and net benefit

#### A.1 Model

For a record i with true class y (relevant or irrelevant), K models cast votes $V_{i1}, \ldots, V_{iK} \in \{0, 1\}$, where 1 means "include". We treat the models as exchangeable draws from a population of models. By de Finetti's theorem (exactly for an unlimited population of models; for a finite population of N models only approximately, with an error of order K/N [S1]) the votes are then conditionally independent given a record-level inclusion probability $p_i$, and we let $p_i \mid y \sim \text{Beta}(a_y, b_y)$. The number of inclusions $S_i = \Sigma_j V_{ij}$ is beta-binomial with mean competence $\bar{s} = a/(a + b)$ and intraclass correlation $\rho = 1/(a + b + 1)$. $\rho = 0$ corresponds to independent errors (the classical Condorcet setting) and $\rho = 1$ to models that always err on the same records. A threshold rule includes a record if $S_i \geq t$: $t = 1$ is "any model includes", $t = \lceil K/2 \rceil$ is majority with ties sent to inclusion, and $t = K$ is unanimity.

#### A.2 Result (i): effective number of models

The variance of the mean vote within a class is $\bar{s}(1 - \bar{s})[1 + (K - 1)\rho]/K$, so K correlated models carry the information of $K_{eff} = K/[1 + (K - 1)\rho]$ independent ones (the design effect). As K grows, $K_{eff}$ increases to $1/\rho$ and never exceeds it.

#### A.3 Result (ii): misses under "any model includes"

A relevant record is missed only if all K models exclude it: $P(\text{miss}) = E[(1 - p)^K] = B(a_1, b_1 + K)/B(a_1, b_1) = \Gamma(a_1 + b_1)\Gamma(b_1 + K)/[\Gamma(b_1)\Gamma(a_1 + b_1 + K)]$. As K grows, $\Gamma(b_1 + K)/\Gamma(a_1 + b_1 + K)$ behaves as $K^{-a_1}$, so $P(\text{miss}) \approx [\Gamma(a_1 + b_1)/\Gamma(b_1)]\ K^{-a_1}$: a power law, not the geometric decay $(1 - \bar{s})^K$ of independent models. With the calibration parameters below, the constant is 0.107 and the exponent −0.553, that is, about $10.7\ K^{-0.55}$ missed records per 100 relevant.

#### A.4 Even majorities equal odd ones

Take 2m exchangeable votes with total T and break ties at random. The majority is correct with probability $P(T \geq m + 1) + \tfrac{1}{2}P(T = m)$. Remove one vote at random. If $T \geq m + 1$ the remaining $2m - 1$ votes still have a majority; if $T = m$ the removed vote is an inclusion with probability ½, leaving $m - 1$ (no majority) or m (majority); if $T \leq m - 1$ there is no majority. The majority of the remaining $2m - 1$ votes is therefore correct with probability $P(T \geq m + 1) + \tfrac{1}{2}P(T = m)$. By exchangeability, a random subset of $2m - 1$ voters has the same distribution as any fixed one, so a majority of 2m with random ties is exactly as accurate as a majority of $2m - 1$. In the calibration corpus the numerical difference was $4 \times 10^{-15}$.

#### A.5 Result (iii): two voters and an adjudicator

If the first two votes agree, the majority of three decides as they do whatever the third vote; if they disagree, the third vote decides. Calling the third model only on disagreements therefore reproduces the majority of three record by record (0 discordant records in the calibration corpus), with expected calls per record of $2 + 2\mu(1 - \mu)(1 - \rho)$, where $\mu$ is the mean inclusion probability in the class. Under the same model, as K grows a majority includes a record only if $p_i > ½$, so its sensitivity tends to $P(p > ½) = 1 -$

$I_{½}(a_1, b_1)$. For odd K (and, by A.4, even K with random ties) it stays below this limit whenever $a_1 > b_1$, because sensitivity minus the limit equals the integral over $p < ½$ of P(majority | p)[f(p) − f(1 − p)], and $f(p) < f(1 − p)$ for $p < ½$. With ties sent to inclusion, an even majority can exceed it.

#### A.6 Calibration corpus and fit

The calibration corpus had 100 records from a review of biologics for severe asthma, 21 of them included by human reviewers, screened with a pre-specified protocol. Ten configurations from two developers, mostly of earlier model versions, voted on every record: Claude Haiku 4.5, Sonnet 5 and Opus 5 through the API; the same three as agents; Gemini 3.5 Flash-Lite (the version used later in the study), Gemma 4 26B and Gemma 4 31B through the Google API; and one further independent Claude run. Seven of the ten were Claude configurations, and repeated runs of one model are highly correlated (intraclass correlation 0.91), which probably raises the estimated ρ. Six of 1,000 votes were missing and counted as "include". Maximum likelihood by class gave $a_1 = 0.553$, $b_1 = 0.0716$ for relevant records ($\bar{s} = 0.885$, $\rho = 0.615$) and $a_0 = 0.234$, $b_0 = 0.594$ for irrelevant records (mean inclusion 0.282, $\rho = 0.547$). The Fleiss intraclass correlation, a different estimator, was 0.63 on relevant records (95% CI 0.35–0.82). The model underestimated the mean misses over all subsets for "any model includes" with one to three models, matched them at four (predicted versus observed misses per 100: 11.5 vs 13.8 with one model, 7.6 vs 9.4 with two, 6.0 vs 6.7 with three, 5.1 vs 4.9 with four) and over-predicted them beyond (3.0 vs 0.0 with ten), because 21 relevant records cannot show a tail. It overestimated majority sensitivity by 4–6 percentage points (three models: 0.893 predicted vs 0.852 observed), because real models differ in competence. Mean absolute error between predicted and observed sensitivity and specificity was 0.028 (60 comparisons).

#### A.7 Net benefit and the value of a study

Net benefit per screened record, in human readings, is NB = $\pi \cdot$ sensitivity $\cdot$ V/h − $(1 − \pi) \cdot$ FPR − machine cost/h. V/h is normative, but the double human screening standard reveals a lower bound: if a second human reviewer, costing one reading per record, is justified even though it only recovers Δs of the relevant studies, the standard values a study at $V/h \geq 1/(\pi \cdot \Delta s)$ readings, with $\Delta s = m(1 − m)(1 − \rho_h)$. With a single-reviewer miss rate m of 10%–13% (10.5% for a pharmacological topic and 13.4% overall in a randomised trial [2]), human–human error correlation $\rho_h$ of 0–0.3 and prevalence of 1%–5%, this bound runs from about 180 to about 1,600 readings per study (about 140 with the 16.8% miss rate the same trial observed for a public health topic). One human reading was costed at 38 seconds (reported by reviewers, not timed) at US$25 per hour, US$0.264.

#### A.8 Optimal size

K95 is the smallest number of models whose best configuration (over all subsets of that size and both rules) reaches 95% of the maximum net-benefit gain over the best single model. It was computed in each bootstrap replicate, resampling reviews and then records within reviews, for each of 12 scenarios (π of 2%, 5% and 10% × V/h of 100, 250, 500 and 1,000). A flat optimum makes the exact K hard to estimate but cheap to miss by one; the calibration corpus alone could only bound K from above. No formal sample-size calculation was made: sizes followed the selection rules in Appendix B.1 and the available budget.

### Appendix B. Data sets, models and settings

#### B.1 Data sets

**Derivation (SYNERGY v1.0).** Fourteen clinical reviews in humans with recoverable criteria: Appenzeller-Herzog 2019, Bos 2018, Brouwer 2019, Chou 2003, Chou 2004, Donners 2021, Jeyaraman 2020, Meijboom 2021, Nelson 2002, Oud 2018, van Dis 2020, Wolters 2018, van der Valk 2021 and van der Waal 2022. For each review, all relevant records (final inclusion) up to 45 and four irrelevant records per relevant one were sampled (seed 20261020; 2,045 records, 409 relevant; abstracts rebuilt from OpenAlex). Deduplication by the screening software removed nine irrelevant records. The paired sample took every relevant record and an equal number of irrelevant ones per review (seed 20261021; 814 records). The primary analysis excluded Jeyaraman 2020 (Appendix C): 13 reviews, 724 records, 364 relevant and 360 irrelevant. Criteria came from the full-text methods section in eight primary reviews (266 relevant records) and from the abstract in five without accessible full text (Bos 2018, Wolters 2018, Chou 2003, Chou 2004, Nelson 2002; 98 relevant records). Haiku and Sonnet first voted as agents on batches of the full sampled corpus (one relevant record per four irrelevant); Opus, Fable, the other seven models, the second agent runs of Haiku and Sonnet and their API runs used batches of the paired sample.

**Replication 1 (CLEF TAR 2019, registered).** Labels from CLEF TAR 2019 Task 2 (test and training topics). Eligible reviews had 10–150 records relevant at title–abstract level; within type they were ordered by the SHA-256 hash of their identifier. DTA: CD009694, CD010657, CD009944, CD012080, CD011548. Intervention: CD011768, CD010778, CD012551, CD006715, CD011977. All title–abstract relevant records up to 45 per review and as many irrelevant records at random (seed 20261025): 740 records, 370 relevant, 194 finally included; 46 records had no abstract and were screened on title. Criteria came from the most recent review version with an abstract (.pub3 for CD012551 and CD006715).

**Replication 2.** Sixteen further reviews by the same rule (relevant records up to 20 per review plus as many irrelevant; seed 20261028): DTA CD009551, CD007431, CD008803, CD011515, CD009372, CD009647, CD009135, CD010864; intervention CD007867, CD001261, CD012347, CD009642, CD009069, CD006468, CD008170, CD010753. 626 records, 313 relevant, 146 finally included; 35 records without abstract. Criteria came from the latest review version published before 2019 (for CD012347, the 2019 version, the earliest available).

**Complete CLEF collections.** Batch 1 (sealed 28 October 2026): collections of 150–450 records with at least five relevant records, prevalence of at most 6% and at least 80% of relevant records with an abstract, not used before, ordered by the SHA-256 hash of their identifier and taken until three DTA and three intervention reviews were filled; six reviews, 1,539 records. Batch 2 (sealed 29 October 2026) used the same rule with collections of 150–1,500 records, three DTA and two intervention reviews and at most 6,000 records, excluding all reviews used before: five collections, 4,380 records. Criteria came from the "Objectives" and "Selection criteria" of each Cochrane abstract. Together: 11 collections (6 DTA, 5 intervention), 5,919 records, 217 relevant at title–abstract level (3.7%), 96 finally included (1.6%), 509 without abstract.

**Reviews published in 2026.** Criteria were written and hashed before searching (30 October 2026): human clinical systematic reviews of randomised trials or diagnostic accuracy studies with an electronic publication date from 1 July to 30 October 2026; no earlier version; a CC BY or CC BY-NC licence; a complete MEDLINE strategy with a search date that could be rebuilt in PubMed with a count within ±15% of the reported one; an included-study list mappable to PMIDs with at least five retrieved by the rebuilt search; 200–4,000 records; and eligibility criteria stated in the abstract. Of 42 candidates, 33 were verified (nine, flagged by the searchers as probable exclusions, were not); six were eligible and all six were taken: CD016389 (home visits after preterm birth), PM42877099 (preterm birth with a short cervix), PM42532626 (augmentation in major depression), PM42688617 (GLP-1-based drugs for weight loss), PM42775083 (fetal lung Doppler) and PM42386350 (splenic stiffness for varices in children). The collections were the complete rebuilt PubMed results (6,277 records; 208 without abstract). Two independent LLM agents (Claude, Anthropic), each blind to the other's output, mapped the included studies to PMIDs from the article, its supplementary material and PubMed (209 of 210 PMIDs agreed; two citation errors were corrected and one erratum notice removed). The reference standard counts PubMed records: the 223 included studies had 208 records (some several, some none); 178 were in the collections and 30 more lay outside the rebuilt searches and were not counted. In CD016389, the 36 records found are reports of 20 of its 35 included studies. An LLM agent transcribed the six eligibility protocols blind to included-study lists and corpora. The documented training cut-off of Claude Haiku 5.5 is June 2026 [29]. DeepSeek does not publish cut-offs; DeepSeek V4 Pro (released 12 August 2026) and V4.1 Flash (released 10 September 2026; snapshot of 17 October) were released after two and three of the six reviews were published, respectively; the V4.1 Flash snapshot used (17 October) postdates all six (published 1 July–12 October), so only the memory test (Appendix E.7) addresses its contamination. For CD016389, a review by other authors, posted as a preprint in 2022 and published in February 2026 [S2], includes about a third of its included studies (12 of 35; 9 in the preprint).

#### B.2 Models and routes

**Table B.1.** Models, developers, identifiers and calling routes.

| Model | Developer | Tier | Identifier and route | Stages |
|---|---|---|---|---|
| Claude Haiku 5.5 | Anthropic | Mid | Agent in Claude Code (derivation, replication 1); claude-haiku-5-5 via Message Batches API (route control on derivation and replication 1 batches; all stages from replication 2) | Derivation onwards (pipeline voter) |
| Claude Sonnet 5.5 | Anthropic | Mid | Agent; claude-sonnet-5-5 via Message Batches API (route control on derivation batches; replication 2) | Derivation, replications |
| Claude Opus 5.5 | Anthropic | Frontier | Agent in Claude Code | Derivation, replication 1 |
| Claude Fable 5.1 | Anthropic | Frontier | Agent in Claude Code | Derivation, replication 1 |
| Gemini 3.5 Flash-Lite | Google | Mid | OpenRouter, Google AI Studio | Calibration, derivation, replications |
| Gemini 3.8 Flash | Google | Mid | OpenRouter, Google AI Studio | Derivation, replication 1 |
| Gemini 4 Argon | Google | Frontier | OpenRouter, Google | Derivation, replication 1 |
| GPT-6 Luna | OpenAI | Mid | OpenRouter, OpenAI | Derivation, replications |

| Model | Developer | Tier | Identifier and route | Stages |
|---|---|---|---|---|
| GPT-6 Sol | OpenAI | Mid | OpenRouter, OpenAI | Derivation, replication 1 |
| DeepSeek V4.1 Flash | DeepSeek | Mid | deepseek-v4.1-flash-1017 via OpenRouter, Relace | Derivation onwards (pipeline voter) |
| DeepSeek V4 Pro | DeepSeek | Frontier | deepseek-v4-pro-0813 via OpenRouter, Ionstream | Derivation onwards (pipeline adjudicator) |

Settings. OpenRouter calls requested JSON output, temperature 0 (not supported by the OpenAI models), low reasoning effort for the Gemini Flash models and GPT-6 models, medium for Gemini 4 Argon and none for DeepSeek, no fallback and quantisation bf16, fp16, fp32 or unreported (the DeepSeek V4.1 Flash provider later reported fp4); each model was served by one provider throughout, pinned from replication 2 onwards. Anthropic API calls used low effort, a strict JSON schema and default temperature. The sampling settings of the Claude Code agents were not logged; their votes were cast between 21 and 26 October 2026 (UTC). Maximum output length was 32,000 tokens for Anthropic API calls; from replication 2 onwards, OpenRouter calls were capped per batch at 1,200 (DeepSeek V4.1 Flash), 800 (DeepSeek V4 Pro) or 400 (Gemini 3.5 Flash-Lite, GPT-6 Luna) tokens per record plus 4,000, whereas derivation and replication 1 calls had no cap; no seed was set. All models were used as released, without fine-tuning. OpenAI's largest model was excluded for cost, and one further model was dropped after a pilot for cost and latency. API calls were made between 24 October and 4 November 2026.

Prompt. All models received the same English system prompt, a Spanish user instruction (asking for justifications in Spanish), the review's criteria (written in Spanish in a fixed template by LLM agents blind to labels and votes: population, intervention or index test, comparator or reference standard, eligible and ineligible designs, and edge rules) and a batch of up to 50 records (title and abstract, in English). The system prompt was the screening prompt of the authors' pipeline (last modified 12 September 2026), not tuned on derivation or later data and identical in every batch from the derivation onwards; the calibration votes (August 2026) came from earlier pipeline runs and may have used an earlier version. Its full text is in the data deposit. For each record the model returned a judgement per criterion (met, not met or unclear, with a quoted piece of evidence), a decision, a justification of at most 30 words and a confidence. The sensitivity instruction (labelled "balanced" in the code) told models that missing or vague information on population, intervention or comparator counts as met when the design is eligible, and that doubt means include. Gemini 3.5 Flash-Lite often omitted record identifiers; when its number of answers equalled the number of records, answers were aligned by position after checking each justification against its record.

Cost. OpenRouter models were costed at the amount billed; Anthropic API calls from logged tokens at list price, halved for the Message Batches API; Anthropic models voting as agents at list price applied to tokens measured for the other models (later API runs used more tokens per record, so these costs are probably underestimated). Logged API spend for this study from 24 October to 4 November 2026 was US$27.14 on OpenRouter (excluding US$2.82 for pilot runs on other reviews) and US$6.20 on the Anthropic API; earlier calibration runs are not included.

### Appendix C. Pre-specification, seals and deviations

#### C.1 What was fixed before each stage

**Table C.1.** What was fixed before each stage, with seal times (UTC) and hash prefixes.

| Stage | Fixed before any vote (or, where stated, before analysis) | Time (UTC) | Hash (prefix) |
|---|---|---|---|
| Derivation | Corpus and protocols frozen; analysis plan not registered | Before screening | — |
| Replication 1 | OSF registration osf.io/zakg9 (7 predictions) | 25 Oct 2026, 18:14 | Predictions 77ca5e24 |
| Role agents | Protocol, inputs and thresholds (two of three copy runs and a 158-record pilot of two role prompts, excluded from the confirmatory analysis, preceded the seal) | 28 Oct 2026, 20:06 | Set 37b6481d |
| Replication 2 and CLEF batch 1 | Predictions R1–R5, V1–V3; pipeline configuration | 28 Oct 2026, 20:11 | Predictions d0da24a8 |
| CLEF batch 2 and context control | Predictions W1–W3, X1–X2 (written after batch 1 results were known; title–abstract floor lowered from 80% to 70%) | 29 Oct 2026, 17:40 | Predictions 5bc7bb0b |
| Reviews of 2026 | Selection criteria; predictions Z1–Z3; addendum | 30 Oct 2026, 20:15; 1 Nov 2026, 15:05 and 16:06 | 32431f41; 590612f3; c52fdc68 |
| Methods-section criteria | Predictions N1–N3, C | 3 Nov 2026, 14:14 | Seal f4c2ac2e |
| Re-vote, retrospective | Predictions RV1–RV5 and decision table (before analysis of existing votes; design informed by DeepSeek V4.1 Flash runs) | 4 Nov 2026, 13:36 | Seal 8642ac2f |
| Re-vote, prospective | Predictions RP1–RP3 | 4 Nov 2026, 16:22 | Seal e562f457 |

All seals except the OSF registration are local SHA-256 seals without an external time-stamp; the seal files and the sealed documents and scripts are deposited with the data, except sealed files that contain titles and abstracts, withheld labels or raw model outputs (votes are deposited as tables) and internal working documents of the role-agent stage; their hashes remain in the seal files. The OSF registration (osf.io/zakg9) was embargoed at registration and will be made public before submission. A third replication on 40 further CLEF reviews was sealed with batch 2; one of its six models had not voted at the time of writing, and its results have not been examined. A blinded human adjudication of a sample of the pipeline's decisions was sealed on 6 November 2026 (seal 9683dd63) for a separate study and is not reported here; the reference standard of this study is the published reviews' decisions.

#### C.2 Deviations

**Derivation.** (1) Amendment 1, defining an incoherent reference standard, was written after Haiku's votes on Jeyaraman 2020 showed 8 of 45 relevant records found; a random sample of 10 missed relevant records, checked against the protocol without votes, showed that all 10 violated an explicit criterion (six did not use mesenchymal stem cells; four were not randomised trials), above the threshold of five. No other review met the first condition (next lowest, 56%). (2) The unregistered draft protocol (v0.1, 19 October 2026) required criteria from the methods section; five reviews without accessible full text used the abstract. It also planned eight models from three developers, a cap of 40 relevant records, $K \leq 8$, an equivalence test for even versus odd majorities and human adjudication of records contradicted by most models; the study used 11 models from four developers, a cap of 45, K ≤ 5 and bootstrap, without the equivalence test or the adjudication. (3) Haiku and Sonnet voted on batches of the full sampled corpus (one relevant per four irrelevant records); the other models on batches of the paired sample. (4) Fourteen DeepSeek V4.1 Flash decisions remained missing because a resubmission overwrote a more complete first response; they counted as "include". (5) Majority with random ties was evaluated only with the four Anthropic models.

**Replication 1.** (1) Criteria were transcribed from the "Objectives" and "Selection criteria" of each Cochrane abstract, because full texts were not openly available; this was documented before any vote. (2) Four batches were resubmitted once. (3) After registration, two further models (GPT-6 Luna and DeepSeek V4.1 Flash) were repeated; they also agreed more with themselves than with other models (8 of 8; kappa 0.77–0.97), but the registered result is 6 of 6. (4) DeepSeek V4 Pro and Gemini 3.5 Flash-Lite tied as adjudicators (329 of 370). The registration specified no tie-break; the analysis script named DeepSeek V4 Pro by a cost computed with a cheapest-first calling order rather than the registered one (Haiku and Sonnet vote, the third adjudicates), under which Flash-Lite is marginally cheaper (about US$3.01 vs 3.03 per 1,000). P5 is supported with either.

**Replication 2 and CLEF collections.** (1) Four OpenRouter calls in batch 1 were logged at zero cost (about US$0.08). (2) Spending caps were updated 38 seconds after the batch 2 seal; no vote was affected. (3) One DeepSeek V4.1 Flash batch in batch 2 failed twice by exhausting the output limit; its votes counted as missing and the records went to the adjudicator, as the rules required.

**Reviews of 2026.** (1) Before voting, the search date limit of PM42877099 was moved to its 3 February 2026 update, and an addendum kept PM42775083 (implicit population and reference standard) and excluded a borderline review (CD015156.pub2) whose mechanically rebuilt search returned 198 records, below the minimum of 200. (2) One Haiku batch could not be resubmitted (one missing vote) and two DeepSeek V4.1 Flash batches exhausted the output limit; missing votes went to the adjudicator. (3) The DeepSeek V4.1 Flash provider began reporting fp4 quantisation; a wrapper that accepted it reproduced 100 of 100 earlier votes. (4) In the memory test, calls rejected with HTTP 404 before reaching a model were not counted as attempts, contrary to the sealed rule; under that rule all six reviews would have been flagged for a sensitivity analysis.

**Methods-section criteria.** Contrary to the sealed rule, the analysis ran although the spending cap stopped one resubmission of the control arm; the 17 missing control votes included no target or included record.

**Re-vote, prospective.** One batch of 42 records failed three times by exhausting the output limit (11.4% of votes missing), which made all predictions indeterminate by the sealed rule; the replicate pass was stopped by the spending cap.

**Role agents.** Six smoke-test calls ran 8–25 seconds after the seal; their outputs were never read. Arms with four to seven copies and with debate or specialist agents were not run; the sealed code omitted the "reference-dependent" label and two secondary analyses, which the report applied by the sealed criteria; the 1-pp materiality threshold awaited confirmation at sealing.

### Appendix D. The pipeline at real prevalence, by review

**Table D.1.** Included studies reaching human reading and reading load, by review.

| Set | Review | Topic | Type | Records | Pipeline: title–abstract relevant | Pipeline: included; load | Haiku alone: included; load | Either voter: included; load |
|---|---|---|---|---|---|---|---|---|
| CLEF | CD007868 | Fluoride toothpastes for preventing dental caries | INT | 300 | 5/5 | 3/3; 3.3% | 3/3; 7.3% | 3/3; 7.7% |
| CLEF | CD010542 | Transient elastography in alcoholic liver disease | DTA | 348 | 15/20 | 7/8; 7.5% | 7/8; 8.3% | 7/8; 10.1% |
| CLEF | CD004414 | Preventing occupational irritant hand dermatitis | INT | 336 | 12/16 | 7/7; 8.3% | 7/7; 9.2% | 7/7; 9.5% |
| CLEF | CD012120 | Exercise training in spinal muscular atrophy type 3 | INT | 169 | 0/7 | —; 4.1% | —; 1.2% | —; 4.1% |
| CLEF | CD012216 | Florbetapir PET in mild cognitive impairment | DTA | 217 | 5/11 | 1/1; 5.5% | 1/1; 4.6% | 1/1; 5.5% |
| CLEF | CD010896 | Cerebral blood flow SPECT for frontotemporal dementia | DTA | 169 | 5/6 | 3/3; 16.6% | 3/3; 9.5% | 3/3; 16.6% |
| CLEF | CD008892 | Rapid diagnostic tests for enteric fever | DTA | 1,499 | 67/69 | 30/30; 23.1% | 30/30; 20.9% | 30/30; 23.4% |
| CLEF | CD011912 | Pulse oximetry for critical congenital heart defects | DTA | 1,406 | 35/36 | 18/18; 9.0% | 18/18; 7.6% | 18/18; 12.2% |
| CLEF | CD008081 | OCT for diabetic macular oedema | DTA | 970 | 13/26 | 9/10; 6.1% | 9/10; 3.6% | 9/10; 6.3% |
| CLEF | CD000996 | Inhaled corticosteroids for bronchiectasis | INT | 281 | 6/9 | 6/6; 8.9% | 6/6; 5.7% | 6/6; 10.0% |
| CLEF | CD010239 | Oxygen concentration for resuscitating preterm infants | INT | 224 | 10/12 | 10/10; 7.6% | 10/10; 22.8% | 10/10; 22.8% |
| 2026 | CD016389 | Home visits after preterm birth (Cochrane) | INT | 776 | — | 36/36; 14.7% | 36/36; 18.2% | 36/36; 18.6% |
| 2026 | PM42877099 | Preventing preterm birth with a short cervix | INT | 962 | — | 21/26; 10.1% | 23/26; 15.6% | 23/26; 16.6% |
| 2026 | PM42532626 | Augmentation strategies in major depression | INT | 301 | — | 33/37; 27.2% | 33/37; 28.9% | 33/37; 28.9% |
| 2026 | PM42688617 | GLP-1-based drugs for weight loss | INT | 3,195 | — | 52/56; 12.2% | 51/56; 11.8% | 52/56; 14.1% |
| 2026 | PM42775083 | Fetal lung Doppler for neonatal respiratory distress | DTA | 650 | — | 9/9; 12.3% | 9/9; 3.4% | 9/9; 12.3% |
| 2026 | PM42386350 | Splenic stiffness for varices in children | DTA | 393 | — | 12/14; 7.1% | 13/14; 12.7% | 13/14; 14.2% |

Pipeline: Claude Haiku 5.5 and DeepSeek V4.1 Flash vote on every record; DeepSeek V4 Pro adjudicates disagreements and records with one missing vote. Load: proportion of the collection's records sent to human reading. DTA, diagnostic test accuracy; INT, intervention; PET, positron emission tomography;

SPECT, single-photon emission computed tomography; OCT, optical coherence tomography; GLP-1, glucagon-like peptide-1. The 2026 reviews have no title–abstract labels. CD012120 had no finally included study. In PM42775083 one DeepSeek V4.1 Flash batch returned no votes, so its records went to the adjudicator; without that batch the pipeline's load in this review would have been 4.6%.

**Operational figures.** Across the 17 reviews, 742 records (6.1%) went to the adjudicator: 579 (4.7%) because the two voters disagreed and the rest because one vote was missing. DeepSeek V4.1 Flash left 57 of 4,380 votes missing in CLEF batch 2 and 105 of 6,277 in the 2026 reviews, mostly in batches that exhausted the output limit; Haiku left one missing. DeepSeek V4.1 Flash was not deterministic at temperature 0: in resubmitted batches, 10 of 237 votes (4.2%) changed. As adjudicator, DeepSeek V4 Pro included 48.5% of the records it saw in the 2026 reviews and excluded three of the five included studies that reached it; in CD008892 (rapid tests for enteric fever, the CLEF collection with the highest load) it included 144 of 149 disagreements.

**Missed studies, CLEF collections.** Of 217 title–abstract relevant records, 44 did not reach reading; in all of them both voters excluded the record, and 42 of the 44 were excluded by the review authors after reading the full text. A post hoc, unblinded reading by an LLM (Claude, Anthropic) classified one as a clear model error, 10 as debatable and 33 as not meeting the published criteria. Two finally included studies were missed: PMID 18691352 (CD010542; the model read "participants with viral hepatitis were excluded" as a reason to exclude the study, a clear error) and PMID 20530184 (CD008081, debatable).

**Missed studies, 2026 reviews.** Fifteen included studies did not reach reading, all in the five non-Cochrane reviews. Two were clear model errors (a trial with a liraglutide versus placebo arm that no voter noticed; a record for which Haiku marked every criterion as met and still excluded it), one was debatable (progression of varices rather than detection), 11 involved rules that the review applied but did not state in its abstract (trials whose population included a subgroup without diabetes; mixed populations with a previous preterm birth; an olanzapine–fluoxetine combination versus fluoxetine; a wider target condition) and one was a reference-standard error (an open-label study without placebo listed among included trials). Three of the 15 had been included by Haiku and excluded by the adjudicator. This classification was made post hoc, after unblinding, by a single LLM reader (Claude, the same developer as one voter); a blinded human adjudication is planned as a separate study.

### Appendix E. Further analyses

#### E.1 Error correlation by developer and calling route

In the derivation set, mean pairwise kappa on relevant records was: Anthropic pairs 0.64 (6 pairs); Google 0.34 (3); OpenAI 0.52 (1); DeepSeek 0.56 (1); all non-Anthropic within-developer pairs 0.42; between developers 0.41 (44 pairs). The excess agreement within developers therefore came from the Anthropic models, which also shared the agent route. With every model called through the API (replication 2), Haiku and Sonnet still agreed more with each other (kappa 0.69) than models of different developers did on average (0.43), but Haiku agreed as much with DeepSeek V4.1 Flash (0.72), and the two DeepSeek models also agreed more with each other (0.58) than models of different developers did.

Calling route. On identical paired batches of the 724 derivation records, Claude Haiku 5.5 found 317 relevant records in a second agent run and 303 through the API (29 found only as agent, 15 only by API; exact McNemar P = 0.049); Sonnet 5.5 found 320 and 298 (33 vs 11; P = 0.001). Against the first agent runs, which used batches of the full sampled corpus, the differences were similar (Haiku 323 vs 303, P = 0.002; Sonnet 317 vs 298, P = 0.014). Through the API, Haiku was also more specific (FPR 0.107 vs 0.198 in the second agent run on the same batches). The API route also changed reasoning effort and output format. On identical CLEF batches, Haiku found 322 and 319 of 370 (14 vs 11; P = 0.69). Sonnet was not called through the API in CLEF before replication 2.

#### E.2 Repetition of the same model

Each of the six models registered for repetition voted twice on the 814-record paired sample (Haiku's and Sonnet's first runs came from batches of the full sampled corpus); agreement is Cohen's kappa over all records. Kappa between the two runs was 0.80 (Gemini 3.5 Flash-Lite), 0.86 (Haiku), 0.91 (Gemini 3.8 Flash and DeepSeek V4 Pro), 0.93 (Sonnet) and 0.97 (Gemini 4 Argon), against mean kappas of 0.58–0.78 with other models; 12–81 of 814 decisions changed between runs.

#### E.3 Role agents of one model

Seven Claude Haiku 5.5 "agents", each a single API call per batch with no tools or memory (unlike the Claude Code agents of the derivation), voted on 1,554 records (primary set 1,464 records, 734 relevant, 23 reviews from the derivation set and replication 1). One used the standard prompt and six added a role (methodologist, clinician, intervention specialist, protocol auditor, reader starting from the conclusions, information specialist) without changing the criteria. Three identical copies were also compared, and eight different base models served as a descriptive reference. Intraclass correlation on relevant records was 0.78 among copies (95% CI 0.61–0.91), 0.75 among roles (0.62–0.87) and 0.56 among different models (0.44–0.67), so the seven prompts were worth 1.27 independent agents and eight models 1.63. Under "any agent includes", mean sensitivity rose from 0.843 with one agent to 0.892 with three and 0.913 with seven; the gain from three to seven was 2.12 pp (90% CI 1.19–3.58), with the FPR rising from 0.157 to 0.190 (equal weight per review; pooled 0.180 to 0.199). The four extra agents paid for their false positives in 7 of 8 sealed net-benefit scenarios, so the sealed thesis that same-model ensembles saturate at three was not supported, and two structural predictions of the theory (on the order of correlations and on a third copy) were not supported either. Under majority voting, sensitivity did not rise (0.845 with three, 0.837 with seven). Over the best trio, a fourth role agent added 0.68 pp (post hoc). Among the eight different models (descriptive, previously seen data), the best trio chosen within the sample left four or more models material in about 1% of replicates, but a trio chosen on the other reviews and applied to the left-out review fell 2.3–3.4 pp short of all eight, and four or more models were material in 50%–58% of replicates in every scenario. By the sealed rule, the primary verdict was "indeterminate", because a floor test was inconclusive, and "reference-dependent", because against a latent reference the gain fell to 1.47 pp. Spend was US$1.44.

#### E.4 Re-voting exclusions

Retrospectively, using existing repeated runs of seven models on the derivation set, a second call recovered about one in five excluded relevant records. Pooled sensitivity rose from 0.839 with one call to 0.873 when every excluded record was re-voted and records with no unmet criterion were included by rule; most of the gain came from exclusions with a single unmet criterion, which held 73% of excluded relevant records. A post hoc simulation of the pipeline suggested a rise from 85.4%–89.8% to 91.2%–91.5%. Of the five sealed retrospective predictions, three were supported; RV2 (at most 5% recovery of exclusions with two or more unmet criteria; observed 11.4%) and RV4 (more than 180 readings per additional relevant record; observed 33.6) were not. A sealed prospective test, in which DeepSeek V4.1 Flash re-voted its 368 exclusions in replication 1 (47 relevant), was indeterminate: one batch failed three times and 11.4% of votes were missing; among records with a vote, 1 of 43 excluded relevant records was recovered, and the simulated pipeline moved from 91.1% to 91.3%. The gain seen retrospectively for DeepSeek V4.1 Flash came largely from a poor first run.

#### E.5 Methods-section criteria

In three of the 2026 reviews, DeepSeek V4.1 Flash re-voted 1,656 records with the eligibility section of each review's methods copied verbatim, to test whether eight included studies missed with abstract criteria would be recovered. Four of eight were included, against one of eight on a same-day re-vote with the abstract criteria (pre-specified threshold six; not supported); more recoveries than losses (N2) was not supported, and load of at most 20% (N3) was; only the comparator exception in the depression review, which is stated in the methods but not in the abstract, was clearly recovered. Four of the five included studies lost with methods criteria were excluded by the methods text itself. Reading load rose to 16.4%, entirely because 101 votes were missing after outputs exhausted the token limit; among records with a vote it was 11.0% against 11.8% with abstract criteria.

#### E.6 Batch composition (context control)

The same 162 records from batch 2 (81 title–abstract relevant, 81 irrelevant) were screened again in balanced batches. The pipeline's final decision was identical for every relevant record (82.7% in both); two irrelevant decisions changed in opposite directions, one because of a missing vote (4.9% in both). Ten of 324 individual votes changed, most towards inclusion in balanced batches. The test had low power: a difference of 6.9 pp would have been detected only 22% of the time.

#### E.7 Memory test

Asked whether they knew each review and to list its included studies, none of the three pipeline models named any included study of the six 2026 reviews. In the 11 older CLEF reviews, DeepSeek V4.1 Flash named 49 studies (mean recall 24%) and DeepSeek V4 Pro 21 (11%), whereas Haiku claimed to know 10 reviews but named no study, so the test was not informative for Haiku.

#### E.8 Latent class analysis

Without using the published reference, latent class models [S3] of increasing complexity (from conditional independence to random record effects and dependence within developer) estimated the true class from the 11 models' votes in the derivation set and replication 1. Under every latent truth, the best four-model ensemble did not improve on the best trio (0.0 pp), and mixing developers in pairs was not shown to help (+0.54 pp, 95% CI −0.61 to 2.54, in the derivation set; +0.99 pp, −1.26 to 2.99, in replication 1). The latent class was not stably identified: it largely reproduced the vote of one model, which changed between data sets and bootstrap replicates. This analysis was post hoc.

### Appendix F. STARD 2015 checklist (applicable items)

Records stand for participants, the published review's decision for the reference standard, and each model or configuration for the index test.

**Table F.1.** STARD 2015 items and where they are reported.

| Section | Item | Location |
|---|---|---|
| Title or abstract | 1 Identification as a study of accuracy, with at least one measure of accuracy | Title; Abstract |
| Abstract | 2 Structured summary | Abstract |
| Introduction | 3 Scientific background and intended use | Section 1 |
| | 4 Objectives and hypotheses | Section 1; Table 3 |
| Methods: design | 5 Whether data collection was planned before the index test and reference standard were performed | Section 2.1; Appendix C |
| Participants | 6 Eligibility criteria | Section 2.3; Appendix B.1 |
| | 7 On what basis potentially eligible participants were identified | Section 2.3; Appendix B.1 |
| | 8 Where and when potentially eligible participants were identified | Appendix B.1 |
| | 9 Whether participants formed a consecutive, random or convenience series | Section 2.3; Appendix B.1 |
| Test methods | 10a Index test, in sufficient detail to allow replication | Section 2.4; Appendix B.2 |
| | 10b Reference standard | Section 2.3 |
| | 11 Rationale for choosing the reference standard | Sections 2.3, 4.4 |
| | 12a Definition of positivity of the index test (pre-specified or exploratory) | Sections 2.4–2.5 |
| | 12b Definition of positivity of the reference standard | Section 2.3 |
| | 13a Whether index test readers had access to other information | Section 2.1 (blind to reference and other votes) |
| | 13b Whether reference standard assessors had access to index test results | Not applicable (published decisions); Appendix D |
| Analysis | 14 Methods for estimating and comparing accuracy | Sections 2.6–2.7 |
| | 15 Handling of indeterminate results | Section 2.4 (missing decision counted as include) |
| | 16 Handling of missing data | Section 2.4; Appendix D |
| | 17 Analyses of variability (pre-specified or exploratory) | Table 3; Appendix E |
| | 18 Intended sample size and how it was determined | No formal calculation; set by selection rules and budget (Section 2.3; Appendices A.8, B.1) |
| Results: participants | 19 Flow of participants | Fig. 1; Table 1 |
| | 20 Baseline characteristics | Table 1; Appendix B.1 |
| | 21a Distribution of severity of disease | Not applicable |
| | 21b Distribution of alternative diagnoses | Not applicable |
| | 22 Time interval between index test and reference standard | Not applicable |

| Section | Item | Location |
|---|---|---|
| Test results | 23 Cross tabulation of index test by reference standard | Tables 2 and 4; data deposit |
| | 24 Estimates of accuracy and their precision | Tables 2–4; Section 3 |
| | 25 Adverse events | Not applicable |
| Discussion | 26 Limitations | Section 4.4 |
| | 27 Implications for practice | Section 4.3 |
| Other | 28 Registration number and registry | Replication 1: OSF, osf.io/zakg9 (reference 24); later stages locally sealed (Section 2.1; Appendix C) |
| | 29 Where the full protocol can be accessed | Appendix C; data deposit |
| | 30 Sources of funding | Funding statement |

### Appendix G. TRIPOD-LLM checklist (applicable items)

The LLM task is title–abstract screening, a classification task in a study of LLM evaluation (codes E and C in the TRIPOD-LLM checklist [26]); the data are bibliographic records (titles and abstracts) and the ground truth is the published review's decision, so items written for patient or electronic health record data are mapped to their equivalents for records or marked not applicable.

**Table G.1.** TRIPOD-LLM items and where they are reported.

| Section | Item | Location |
|---|---|---|
| Title | 1 Identify the study as evaluating an LLM; task, population and outcome | Title (LLMs, screening task and evaluation named; outcome not named) |
| Abstract | 2 Abstract (TRIPOD-LLM for Abstracts, items 2a–2l) | Abstract |
| | 2a Title: study type, task, population and outcome | Title; Abstract, Background and Objectives |
| | 2b Background: context, use case and rationale | Abstract, Background and Objectives |
| | 2c Objectives: development, tuning and/or evaluation | Abstract, Background and Objectives |
| | 2d Key elements of the study setting | Abstract, Methods |
| | 2e Data used, splits and selective use of data | Abstract, Methods (sampling of records within reviews in Section 2.3) |
| | 2f Name and version of the LLMs | Not reported in Abstract (developers only; names in Table 2) |
| | 2g LLM-building steps | Not applicable: no model built or fine-tuned |
| | 2h Tasks performed by the LLMs; key inputs and outputs | Abstract, Background and Objectives; Methods |
| | 2i Evaluation data, endpoint, hold-out and measures | Abstract, Methods and Results (post-cut-off reviews mentioned only in the Plain Language Summary) |
| | 2j Overall results and interpretation | Abstract, Results and Conclusion |
| | 2k Broader implications or concerns | Abstract, Conclusion |
| | 2l Registration | Not in Abstract (item for healthcare-setting evaluations); OSF registration in Section 2.1 |
| Introduction: background | 3a Healthcare context, use case and rationale; existing approaches | Section 1 |
| | 3b Target population, intended use and intended users | Section 1 (second reviewer, filter or autonomous screener); Section 4.3; What is new? (review teams) |
| Introduction: objectives | 4 Study objectives (development, tuning or evaluation) | Section 1 (last paragraph); Section 2.1 |
| Methods: data | 5a Data sources for training, tuning and evaluation; rationale | Section 2.3; Table 1; Appendices A.6, B.1 (evaluation data only) |

| Section | Item | Location |
| --- | --- | --- |
| | 5b Data points and their distribution; descriptors | Table 1; Section 2.4 (language); Appendix B.1; Table D.1 |
| | 5c Dates of oldest and newest text items | Not reported for records; review publication dates only (Table 1; Appendix B.1) |
| | 5d Pre-processing and quality checking | Section 2.3; Appendix B.1 (abstracts rebuilt from OpenAlex, deduplication, records without abstract) |
| | 5e Missing and imbalanced data; omitted data | Sections 2.3, 2.4 (missing vote counted as include), 2.6 (reweighting of irrelevant records); Appendices B.1, C.2 |
| Methods: analytical methods | 6a LLM name, version and last date of training | Table 2; Appendix B.2 (identifiers for Anthropic API and DeepSeek models only); training cut-off reported only for Claude Haiku 5.5 (Section 2.3; Appendix B.1) |
| | 6b LLM development process (architecture, training, alignment) | Not applicable: existing models used as released, without fine-tuning (Appendix B.2) |
| | 6c Text generation: prompting, consistency and inference settings | Section 2.4; Appendix B.2 (settings, maximum output length from replication 2, no seed; agent settings not logged); consistency in Appendices D and E.2 |
| | 6d Initial and post-processed output | Appendix B.2 (criterion judgements, decision, justification, confidence); Sections 2.4–2.5 (binary vote and aggregation) |
| | 6e Classification rationale; probabilities and thresholds | Sections 2.2, 2.5–2.6; Appendices A.1, A.8, B.2 |
| Methods: LLM output | 7a Quality metrics of outputs against the gold standard | Sections 2.6–2.7; Tables 2–4; error types in Appendix D (item written for generative tasks) |
| | 7b Relevance of metrics to the downstream task | Section 2.6; Appendix A.7 (net benefit in human readings) |
| | 7c Outcome, prediction route, date of inference and metrics | Sections 2.3–2.6; Appendix B.2 (agent votes 21–26 October 2026, UTC; API calls 24 October–4 November 2026) |
| | 7d Subjective assessment: assessors, instructions, agreement | Appendix D (post hoc classification of missed studies by one unblinded LLM reader, Claude); no human adjudication in this study; reference standard = published reviews' decisions (planned separately, Section 4.4) |
| | 7e Comparison with other LLMs, humans and benchmarks | Sections 2.5, 3.2–3.5; Tables 2–4; human miss rates in Section 1 and Appendix A.7 |
| Methods: annotation | 8a How text was labelled; annotation guidelines | Section 2.3 (labels from published reviews); Appendix B.1 (2026 labels from included-study lists mapped to PMIDs; mapping instructions not reported) |
| | 8b Number of annotators, overlap and agreement | Appendix B.1 (two independent PMID mappings, 209 of 210 agreed); Appendix D (one reader) |
| | 8c Background of annotators or models used in labelling | Section 2.3 and Appendix B.1 (LLM agents, Claude, Anthropic, for criteria transcription and PMID mapping); Appendix D (Claude as post hoc reader) |
| Methods: prompting | 9a Prompt design, curation and selection | Final prompt in Section 2.4 and Appendix B.2; design and selection process not reported |
| | 9b Data used to develop the prompts | Appendix B.2 (not tuned on study data) |
| Methods: summarisation | 10 Pre-processing before summarisation | Not applicable: no summarisation task |
| Methods: instruction tuning/alignment | 11 Instructions, data, interface and evaluators | Not applicable: no instruction tuning or alignment |

| Section | Item | Location |
|---|---|---|
| Methods: compute | 12 Compute or proxies (cost, time, machines) | Section 2.6; Tables 2 and 4; Appendices B.2, E.3 (cost; inference time not reported) |
| Methods: ethical approval | 13 Ethics committee approval or waiver | Ethics statement (not required: published records, no human participants) |
| Methods: open science | 14a Funding and role of funders | Funding statement |
| | 14b Conflicts of interest | Declaration of competing interest; Section 4.4 |
| | 14c Access to the study protocol | Section 2.1; Appendix C.1 (sealed files in the data deposit) |
| | 14d Registration | Section 2.1 (OSF osf.io/zakg9, replication 1); Appendix C.1 |
| | 14e Availability of data | Data availability statement (DOI to be added) |
| | 14f Availability of code | Data availability statement (analysis code; DOI to be added) |
| Methods: public involvement | 15 Patient and public involvement | Not applicable: no patients; methodological study |
| Results: participants | 16a Flow of data, with and without the label | Not applicable (no patient data); record flow in Fig. 1, Table 1 and Appendix B.1 |
| | 16b Characteristics by data source and split; key dates | Not applicable (no patient data); data sets in Table 1, Appendix B.1 and Table D.1 |
| | 16c Clinical variables in development and evaluation data | Not applicable: no clinical outcomes |
| | 16d Participants and outcome events in each analysis | Not applicable (no patient data); records and relevant records per analysis in Tables 1, 2 and 4 |
| Results: performance | 17 Performance on pre-specified metrics | Sections 3.1–3.6; Tables 2–4; Figs. 2–5; Appendices D–E |
| Results: LLM updating | 18 Results of LLM updating | Not applicable: no model updated or retrained |
| Discussion: interpretation | 19a Overall interpretation, including fairness | Sections 4.1–4.2 (fairness not discussed) |
| Discussion: limitations | 19b Limitations: biases, uncertainty, generalisability | Section 4.4 |
| Discussion: usability in context | 19c Known challenges of the data (representation, missingness, bias) | Sections 4.3–4.4; Appendices B.1, D |
| | 19d Intended use: input, end-user, autonomy and oversight | Sections 2.6, 3.5, 4.3 (implicit: records excluded by the models are not read); not stated explicitly |
| | 19e Handling of poor-quality or unavailable input | Sections 2.4 and 4.3 (missing votes never count as exclusions); Appendix B.1 (records without abstract screened on title) |
| | 19f User interaction and expertise required | Sections 2.3, 4.3 (criteria template; local validation); expertise not stated |
| | 19g Next steps for research | Sections 4.3–4.4 |

Item wording is abbreviated from the published checklist [26]; item 2 refers to TRIPOD-LLM for Abstracts, whose items 2a–2l are listed. LLM, large language model; OSF, Open Science Framework; PMID, PubMed identifier.

### Supplementary references

[S1] Diaconis P, Freedman D. Finite exchangeable sequences. Ann Probab 1980;8:745-64. https://doi.org/10.1214/aop/1176994663.

[S2] Wilson A, Bedwell C, Actis Danna V, Tate N, Dwan K, Portela A, et al. Interventions to support parents, families and caregivers in caring for preterm or low birth weight infants at home: a systematic review and meta-analysis. PLOS Glob Public Health 2026;6:e0005690. https://doi.org/10.1371/journal.pgph.0005690.

[S3] Qu Y, Tan M, Kutner MH. Random effects models in latent class analysis for evaluating accuracy of diagnostic tests. Biometrics 1996;52:797-810. https://doi.org/10.2307/2533043.
